# Brain Size: To Adjust or Not Adjust? It’s Not a Matter of If, but How

**DOI:** 10.1101/2025.09.21.25336298

**Authors:** Aliza Brzezinski-Rittner, Roqaie Moqadam, Yashar Zeighami, Mahsa Dadar

## Abstract

Total intracranial volume (TIV) is a major confounding factor in neuroimaging studies, particularly when studying sex differences in the brain. Different methods have been proposed to adjust for this effect, however, their impact has not been directly studied and compared. In this study, we sought to evaluate the impact of four most commonly used adjustment methods in the literature on the estimations of neuroanatomical sex differences. These methods included: the proportions method, the residuals method, the power corrected proportions method, and adding TIV as a covariate in a regression analysis. Leveraging data from the UK Biobank, we employed a matching approach to devise a gold standard as reference for comparing these methods. To achieve this, we matched the male and female participants based on age and TIV to remove the impact of TIV differences between sexes. We further modeled aging trajectories at the regional level, vertexwise, and voxelwise, using raw and adjusted values, and compared the obtained estimates against the gold standard. We found that across different metrics, adding TIV as a covariate was the best-performing method for removing the effect of TIV, in terms of the correlation between the estimates of the different subsamples and the gold standard as well as the degree of estimation bias. Furthermore, we showed that the commonly used smoothing of the morphometric measures can result in biased estimation of sex differences in these measures. Finally, we showed that while small in effect size, there still remains some neuroanatomically specific uncorrected effects for all adjustment methods.

## Introduction

Magnetic resonance imaging (MRI) is one of the most commonly used tools for studying structural brain changes. Regional brain volumes have been used in numerous studies, comparing healthy individuals and diseased populations. For example, volumetric studies consistently report smaller hippocampal volumes in patients with Alzheimer’s disease ^1–3^. Another commonly used metric to assess brain changes is cortical thickness (CT), which measures the depth of the cortical layers as the distance between the white matter and gray matter boundaries and the brain’s pial surface ^4^. For example, Parkinson’s disease patients reportedly experience greater levels of cortical thinning compared to matched healthy controls ^5^. Similarly, cortical surface area (SA) can be used as a measure of “cortical column generation” ^6^. Other commonly used metrics to study brain atrophy are deformation based morphometry (DBM), which captures local atrophy at the voxel level by estimating the deformation a preprocessed image goes through in a non-linear registration to an average template ^7^, and voxel based morphometry (VBM), which derives tissue probability maps modulated by DBM as an estimation of density for different tissue types. For example, frontotemporal dementia patients experience widespread atrophy as measured by DBM in frontal and temporal brain regions ^8,9^. Regional volumetric measures in stereotaxic space can be further estimated based on these voxelwise measures. Interestingly, these metrics (volumes, CT, SA, DBM, and VBM) can scale differently in relation to brain size as measured by total intracranial volume (TIV) ^10^.

Imaging-derived findings based on these metrics are inevitably impacted by a series of methodological and processing choices. Of particular interest are the choices made to account for the effect of head size differences when performing population level analyses such as investigating group differences in case-control studies. For example, it has been shown that brain size impacts brain asymmetry findings, and that appropriate adjustment of this effect reveals specific age and sex effects on brain asymmetry ^11^. Another methodological choice pertains to smoothing (at vertex or voxel level), which is commonly used to improve registration and inter-subject alignment, reduce noise, and increase statistical power^12–16^ . However, when it comes to regional analyses, the smoothing level and resolution of the selected parcelation might have a differential effect on the results ^13^.

It has been shown that many of the reported sex differences in the brain are driven by intracranial volume, which is on average 12% larger in males compared to females ^17,18^. While most cortical and subcortical brain regions appear to have significantly larger volumes in males than in females, females present with greater overall cortical thickness, and the effect of these differences decrease, or reveal different patterns when the analyses appropriately account for intracranial volume ^19,20^. A common metric used for adjustments (also referred to as normalization) is the TIV, as it is assumed to be constant across the individual’s lifespan and is largely unaffected by aging or pathological processes ^21–23^. Furthermore, when it comes to cortical measures such as cortical volume (CV), CT, or SA, researchers have used specific “global metrics” to adjust statistical models for these values, namely total cortical volume, mean CT, or total SA ^24^. However, the impact of these choices have also not been systematically studied.

There have been multiple proposed methods for adjusting for TIV, as well as attempts to evaluate and compare these methods ^23,25,26,26–34^. Most studies have focused on a small number of brain structures, instead of taking a brainwide approach. Furthermore, the vast majority of these studies centered exclusively on volumetric or VBM data, and few have used an approach that permits comparisons to a gold standard (i.e., matching). More recently, studies have utilized correction approaches to account for the confounding effect of brain size on sex differences, with a focus on accurate classification of females vs males based on structural MRI measures (beyond brain size) ^31,35^. While employing correction methods removes (some) of the TIV-related differences, each method might also introduce different types of bias when assessing the potential presence of sex differences in normalized values, and in the absence of a gold standard approach, the results remain inconclusive.

Brain size correction methods have been mainly studied in the context of regional brain volumes. Among these, one of the first to be proposed was the *proportions* method, which consists of dividing any given volume of interest (VOI) by TIV, and assumes a proportional relationship between any VOI and TIV, which contradicts the allometry principle in brain development^36–39^. Later, the *residuals* method was proposed^40^ by Arndt et al. 1991, ^41^ and later re-evaluated and expanded by Mathalon et al. 1993 ^40^, which “involved performing a linear regression of each of the raw measures on head size within the normal control group and then using the residuals from this ‘control regression line’ as the head-size-corrected unit of analysis for each participant (i.e., control participant and patients).” ^40^, thus expressing the adjusted VOI as deviation from the predicted value in a healthy participant with a given TIV. While accounting for potential regional differences, an underlying assumption of this method is a linear relationship between VOIs and TIV. The residuals method was developed to examine group differences between diseased and healthy populations, hence it is based on the idea of using the residual values from the control group to correct both groups. For other comparisons (e.g. sex differences in healthy populations), the residuals are calculated using the whole sample ^30^. The *power-corrected-proportions* (PCP) method ^42^ was developed with the idea that the relationship between several VOIs and TIV follows the power law principle ^10,43^, instead of being either proportional or linear. According to this principle, VOI and TIV are related through 𝑉𝑂𝐼 = α𝐼𝐶𝑉^β^ , and one can use an estimation of β to account for this power relationship when using the proportion of VOI to TIV. Lastly, a common practice in neuroimaging studies is adding TIV as a covariate in statistical models, or regressing it out with a simple linear regression before performing any additional analyses ^44–50^, which similar to the regression method, while accounting for regional differences, is based on the same assumption of linearity.

In this work, we aim to a) evaluate the effectiveness of different brain size adjustment methods in removing the effect of brain size on sex differences in aging trajectories using different structural imaging metrics, b) identify the remaining bias after applying different correction methods, and c) assess the effect of smoothing levels on derived sex differences for different adjustment methods. To address these questions, we used a matching approach similar to the one used previously by ^19,29,30,34,51–55^, taking advantage of the large sample size provided by the UK Biobank (UKBB) dataset.

## Methods

### Datasets

Data included participants from the UK Biobank, an open-access large prospective study with phenotypic, genotypic, and neuroimaging data from 500,000 individuals recruited between 2006 and 2010 at 40–69 years old in Great Britain ^56–58^. All participants provided informed consent (“Resources tab” at https://biobank.ctsu.ox.ac.uk/crystal/field.cgi?id=200). The UK Biobank received ethical approval from the Research Ethics Committee (reference 11/NW/0382), and the present study was conducted based on application 45551. Sex was assigned according to the “Sex” variable (data field 31). Brain imaging data was acquired from a subset of participants.

### Image processing

Data included in this study was obtained from the first imaging visit (instance 2). We extracted regional and vertexwise volume, cortical thickness, and surface area information by processing the raw T1-weighted images using FreeSurfer v.7.4.1. ^59^. We used the Desikan-Killiany parcellation ^60^ and aseg segmentations for the cortical and subcortical regions, respectively. Furthermore, we used PELICAN^61^, an extensively validated and widely used ^8,62–65^ in-house pipeline based on the open source MINC and ANTs tools to obtain accurate TIV estimations as well as regional and voxelwise DBM values to capture local atrophy (tissue shrinkage) or expansions ^66–72^. We used PELICAN-derived TIV values as it has been previously shown that the estimated TIV (eTIV) from Freesurfer is biased and might not be optimal for TIV normalization ^26,73,74^. The preprocessing procedure consisted of denoising based on optimized non-local means filtering ^75^, correction for intensity inhomogeneity ^76^, and intensity normalization using linear histogram matching. The resulting images were linearly registered to the MNI152-2009c template (nine-parameters) via a hierarchical linear registration method ^66^. Scaling factors derived from the linear transformations (reflecting the TIV difference between the participant and MNI152-2009c template) were used to estimate TIVs for each participant^61^. Following linear registration, each scan was non-linearly registered to the MNI152-2009c template. We obtained voxelwise DBM maps by using the non-linear transformations to calculate the Jacobian determinant of each participant’s deformation matrix at each voxel. Finally, we also calculated individual regional volumes based on the CerebrA atlas, whose regions are equivalent to the DKT atlas used by FreeSurfer, and further includes subcortical structures ^77^.

Linear and non-linear registrations were visually quality-controlled (QC) by two experienced raters (Y.Z. and M.D.), using a procedure previously described in ^66^. In summary, for the linear registration, QC images were generated by overlaying the contours of the MNI152-2009c template on the registered images. The raters assessed the alignment of the contours with the registered image on sagittal, coronal, and axial views. To assess the alignment, the outline of the brain, central sulcus, cingulate sulcus, and parieto-occipital fissure were used as anatomical landmarks. Ensuring the accuracy of the linear registration process guarantees the accuracy of the scaling factors used for TIV estimation.

### Matching process

Exclusion criteria included: missing demographic information (e.g., age, sex), failed visual quality control of image processing and registration steps, and failure in FreeSurfer’s reconstruction. After these evaluations, our final sample included 35,732 participants (54% females). From this sample, we created five subsamples, which included an equal number of female and male participants, using the same method described in our previous work ^19^. Briefly, the first subset is referred to as the **<u>matched</u> <u>sample</u>,** and was matched by TIV and age, calculated as months between the participant’s month and year of birth (data fields 52 and 34), and the date of visiting the assessment center (data field 53) for the imaging visit (instance 2). The matching process consisted of first splitting the whole sample by sex, randomizing the order within each group, and using an iterative process to find a female whose age in months and estimated TIV were within 0.02% of the corresponding values for a given male. We chose to find females to match to their male counterparts since the female group was larger than the male (19,281 F vs 16,451 M). This process yielded a sample of 11,294 matched participants (5,647 per sex). We created a second subsample with the same size (11,294 participants) matched only by age, referred to as the **<u>age-matched sample</u>**. The third sample we created, referred to as **<u>not-matched sample</u>**, had the same size, however, it mimicked the age and TIV distribution of the original full sample, to allow for comparisons of the results across matching strategies with the same statistical power, and allowing us to investigate the results in a sample characteristic of the complete sample while matching the sample size of the other subsamples. The next sample is referred to as **<u>extreme sample</u>**, in which we exaggerated the TIV difference between females and males by excluding all the participants that were included in the matched sample and keeping 5,647 participants per sex whose age distribution was similar to the original sample. Table 1 summarizes the information for all generated samples.

**Table 1:** Description of the characteristics of the subsamples included in this study.

| Sex | N | Age range | Age (mean, sd) | TIV (mean, sd) |
| --- | --- | --- | --- | --- |
| **Full dataset** |  |  |  |  |
| Female | 19,281 (54%) | 45 - 81 years | 62.8, (7.36) | 1,336,544.0, (103,196.89) |
| Male | 16,451 (46%) | 44 - 82 years | 64.0, (7.64) | 1,525,386.9, (119,264.92) |
| **TIV and age matched sample** |  |  |  |  |
| Female | 5,647 (50%) | 47 - 80 years | 63.3, (7.06) | 1,426,776.1, (83,886.29) |
| Male | 5,647 (50%) | 47 - 80 years | 63.3, (7.06) | 1,426,823.3, (83,880.34) |
| **Age matched sample** |  |  |  |  |
| Female | 5,647 (50%) | 45 - 81 years | 64.0, (7.67) | 1,336,051.7, (103,836.41) |
| Male | 5,647 (50%) | 45 - 81 years | 64.0, (7.67) | 1,525,577.3, (117,849.54) |
| **Not matched sample** |  |  |  |  |
| Female | 5,647 (50%) | 46 - 81 years | 62.8, (7.39) | 1,338,532.7, (103,282.83) |
| Male | 5,647 (50%) | 44 - 81 years | 64.0, (7.67) | 1,524,394.1, (118,165.63) |
| **Extreme sample** |  |  |  |  |
| Female | 5,647 (50%) | 45 - 81 years | 63.6, (7.02) | 1,297,634.6, (85,362.83) |
| Male | 5,647 (50%) | 44 - 82 years | 64.4, (7.95) | 1,574,611.6, (101,046.49) |
| Abbreviations: TIV: Total Intracranial Volume, sd: standard deviation |  |  |  |  |

### Modeling

The modeling strategies described below were used for our eight metrics of interest: vertexwise cortical volume, cortical thickness, and surface area, voxelwise DBM, regional volume, cortical thickness, and surface area, and regional volumes derived from DBM values. Each model was evaluated for each of the five subsamples.

We modeled the aging trajectory of each metric using the following linear regression model (eq. 1), accounting for age, sex, and their interaction:

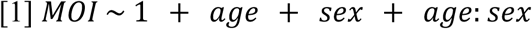

Where *MOI* denotes the measure of interest, *age* is the age at MRI acquisition time in months, and *sex* is the participants’ sex.

To assess whether commonly used adjustment methods can account for head size differences with regard to accurately estimating sex differentiated aging trajectories, we repeated the same aging trajectory analyses (eq. 1) using adjusted values instead of the raw values. An ideal correction method that has appropriately captured the effects of head size differences would yield similar sex differentiated aging trajectories to those obtained based on the matched sample. We examined the following correction methods:

1) Proportions method: for each participant, the regional volume is divided by their TIV, using the following equation:

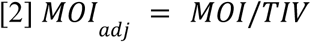

This method has the underlying assumption that all brain VOIs are proportional to TIV.

2) Power-corrected proportions method (PCP) ^42^: the regional volume is adjusted via:

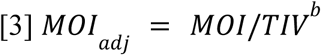

where b represents the slope of the regression between 𝑙𝑜𝑔(𝑀𝑂𝐼) and 𝑙𝑜𝑔(𝑇𝐼𝑉).

3) Residuals method ^40,41^: where the *MOI* is adjusted via the following equation:

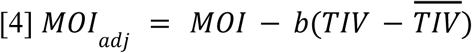

with 𝑏 representing the slope of the regression between 𝑀𝑂𝐼 and 𝑇𝐼𝑉, and 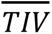 being the mean TIV of the whole sample (following the recommendations by Sanchis-Segura and colleagues (2019) ^30^).

4) TIV as a covariate (or covariate method): we added TIV as a covariate in the regression model, assuming a linear relationship between MOI and TIV:

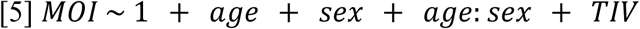

We repeated this procedure in each of the subsamples and compared the model estimates against those obtained based on the raw values using the matched sample.

Furthermore, we examined the impact of different smoothing levels (0, 5, 10, 15, 20, and 25 mm at FWHM) at the vertex level, using vertexwise FreeSurfer outcomes for cortical volumes, CT, and SA. For voxelwise DBM data, we performed the analyses per tissue type. For this, we classified the tissue types based on BISON ^78^ segmentations of the MNI152-2009c template.

Finally, we repeated the analyses replacing TIV with each measure’s “global measure” for adjustment. In other words, we used total cortical volume (TCV) as the adjustment measure for the vertexwise cortical volume analyses, total surface area (TSA) for the vertexwise SA, and mean cortical thickness (MCT) for the vertexwise CT, as employed in some studies using surface-based measurements^20,79,80^. Regional SA and CT were also adjusted using TSA and MCT.

#### Bias estimation

We estimated a bias term for each method and metric, representing the residual bias left for the sex estimation after the corrections were performed for each measure. To achieve this, we compared the local (i.e., regional/vertexwise/voxelwise) sex estimates based on the gold standard matched sample against the corrected estimate in the non-matched sample. In an ideal case, the correction results in an identical estimation to that of the gold standard. We calculated the local estimate differences from this ideal scenario (i.e. the orthogonal distance of each region from the reference line) as the remaining bias in the corrected estimation for each adjustment method. The results were then projected on the brain and visualized to examine whether there is any systematic anatomical pattern to this remaining uncorrected bias.

All statistical analyses were performed using the R statistical language.

## Data and Code Availability Statement

The UK Biobank dataset is open access and can be requested from https://www.ukbiobank.ac.uk/use-our-data/apply-for-access/. FreeSurfer is also open source and freely available at https://surfer.nmr.mgh.harvard.edu/. Similarly, PELICAN ^61^, the image processing pipeline used to derive TIV and DBM measurements is open source and freely available at https://github.com/VANDAlab/Preprocessing_Pipeline.

## Results

### Regional data

#### Regional FreeSurfer volumes

Figure 1a shows the distribution of the standardized model estimates across the 78 brain regions (62 cortical regions from the DKT atlas and 16 subcortical regions from aseg segmentation) for each sample. While age estimations were consistent across the five samples, sex estimations changed between samples due to the TIV differences between females and males. For instance, both non matched and age matched samples had positive sex estimates (with the female group used consistently as reference), indicating that regional volumes were larger for males. The estimations for the extreme sample followed a similar distribution, but were even larger. Meanwhile, in the matched sample, the sex estimates were centered around zero, indicating that although there were some sex differences in the trajectories, they were more subtle and, importantly, bidirectional (i.e. positive for some regions and negative for others).

**Figure 1.**
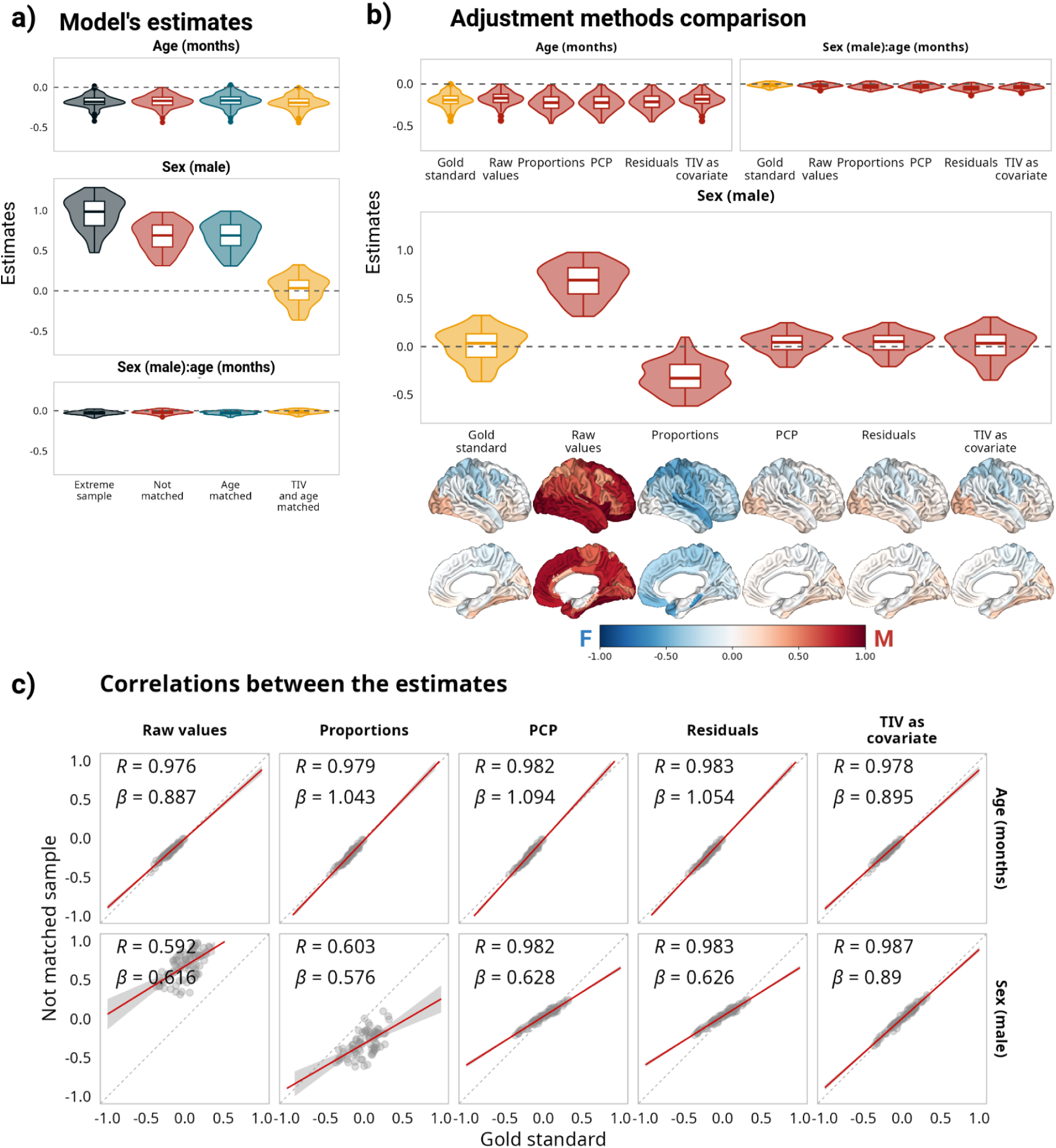
Impact of correction methods on model estimations based on regional volumes. **a**: Distribution of estimates for all samples. **b**: Comparison of estimates for the matched sample without adjustment and the estimates after using different adjustment methods for the not matched sample. Below the sex estimates are the corresponding cortical projections of the same regional values. **c**: Correlations and linear regression slopes between estimates for the matched sample without adjustment and the estimates after using different adjustment methods for the not matched sample. For a comparison of all the estimates, see Supplementary Figure 1.

Figure 1b compares the distribution of the standardized model estimates between the matched and unmatched samples for different correction methods across the same regions and their projection on the brain surface. For the age estimations, while applying correction methods increased their variability, the adjustments had little effect on them (Supplementary Figure 1). For the sex estimates, the proportions method reversed the trend of the not matched sample, and was the only method that significantly changed and reversed the intercept estimations. PCP and residuals methods yielded virtually identical results, both showing the same trend as the gold standard with regards to the sex estimate, while removing some of the variability. Interestingly, this reduction in variability was stronger for the extreme sample, with a systematic bias underestimating larger effect sizes in both directions.

Figure 1c shows the correlation between the model estimates using different adjustment methods in the not matched sample and the gold standard. Both residuals and PCP methods showed high correlations with the gold standard (> 0.98 for intercept, age, and sex estimations). Finally, the model estimates yielded the closest results to the gold standard when TIV was added as a covariate, with an overall distribution that was similar to the estimates of the matched sample without any major bias.

### DBM volumes

Similar to the previous section, Figure 2a shows the distribution of the standardized model estimates across the 78 brain regions for each sample. The age estimates remained constant across the five samples; however, compared to the volumes extracted in the participants’ native space, both the intercepts and the sex estimates were smaller in magnitude and had the opposite direction for the extreme, the non matched, and the age matched samples. This behavior is potentially explained by DBM values already being normalized to a reference template (i.e. MNI-ICBM152) via a linear registration prior to estimating the non-linear transformation. The sex estimates for the matched sample (i.e., gold standard method) were centered around zero and the different adjustment methods (Figure 2b) yielded results equivalent to those obtained with the regional volumes (Figure 1b), with the residuals and PCP methods showing high correlations with the gold standard (ρ = 0.98 for the intercept, age, and sex estimations; Figure 2c). Once again, the model estimates yielded the closest results to the gold standard when TIV was added as a covariate. Interestingly, as opposed to the regional findings, the proportions method did not reverse the trend for the sex estimates but exaggerated the differences. It is worth noting that the normalization that results from linear registration to the ICBM-template is not equivalent to using the proportions method on volumetric information extracted in the participants’ native space (Supplementary Figure 2).

**Figure 2.**
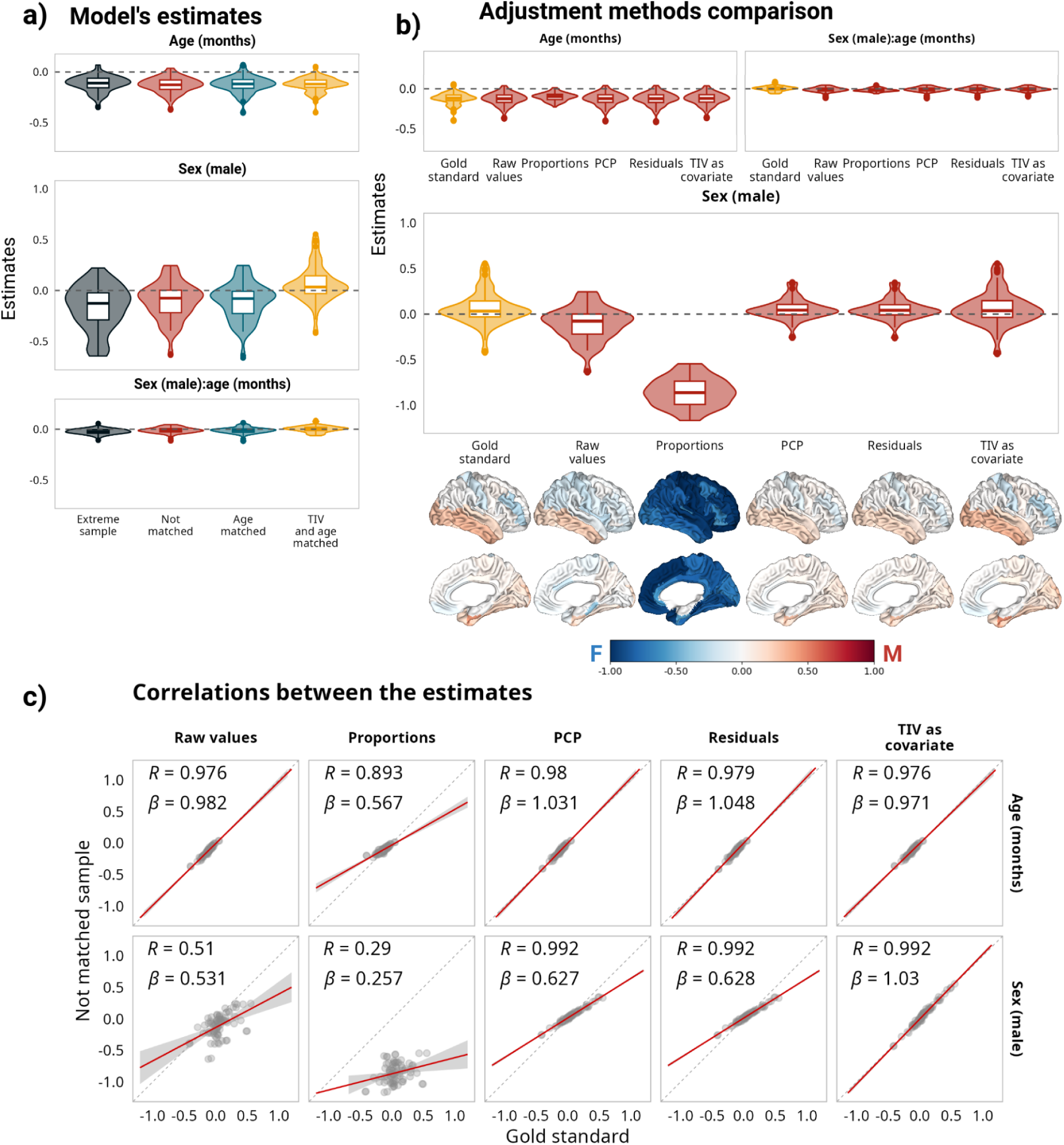
Impact of correction methods on model estimations based on deformation based morphometry (DBM) measures. **a**: Distribution of estimates for all samples. **b**: Comparison of estimates for the matched sample without adjustment and the estimates after using different adjustment methods for the not matched sample. Below the sex estimates are the corresponding cortical projections of the same values. **c**: Correlations and linear regression slopes between estimates for the matched sample without adjustment and the estimates after using different adjustment methods for the not matched sample. For a comparison of all the estimates, see Supplementary Figure 3.

### Regional Surface Area

SA estimates followed a very similar pattern to those of the regional volumes, albeit they only included the 62 cortical regions provided in the DKT atlas. Age was negatively associated with SA for all brain regions and samples. Sex estimations differed across samples, following the same pattern as the regional volumes (Figure 3a). While none of the adjustment methods had a significant effect on the age estimates, in the case of the sex estimates, the proportions method reversed the trends for the non matched, age matched, and extreme sample, with all estimates being negative. PCP, residuals, and adding TIV as a covariate yielded similar results, which were highly correlated with the estimates obtained for the matched sample (Figure 3b). While the correlation values were smaller, the results were also very similar when TSA was used for adjustment instead of TIV (r ∼ 0.9 versus 0.97, respectively; Figure 3d for the adjustment with TIV, and Supplementary Figure 4 for the adjustment with TSA). We further examined the correlation between the estimates obtained while using the two different metrics as adjustment variables. For the age estimate, the correlations for all samples and methods were over ρ = 0.82, with adding the adjusting variable as a covariate showing the highest correlations (ρ = 0.88 for the matched sample, ρ = 0.92 for the not matched, and ρ = 0.94 for the extreme sample; Figure 3e top panel). For the sex estimate, the proportions method had the highest correlation (over ρ = 0.92 for all samples). From the other three methods, adding the adjusting variable as a covariate had the highest correlation in all samples. Interestingly, the correlations for the three methods were lower for the extreme sample and had the highest values for the matched sample (Figure 3e bottom panel).

**Figure 3.**
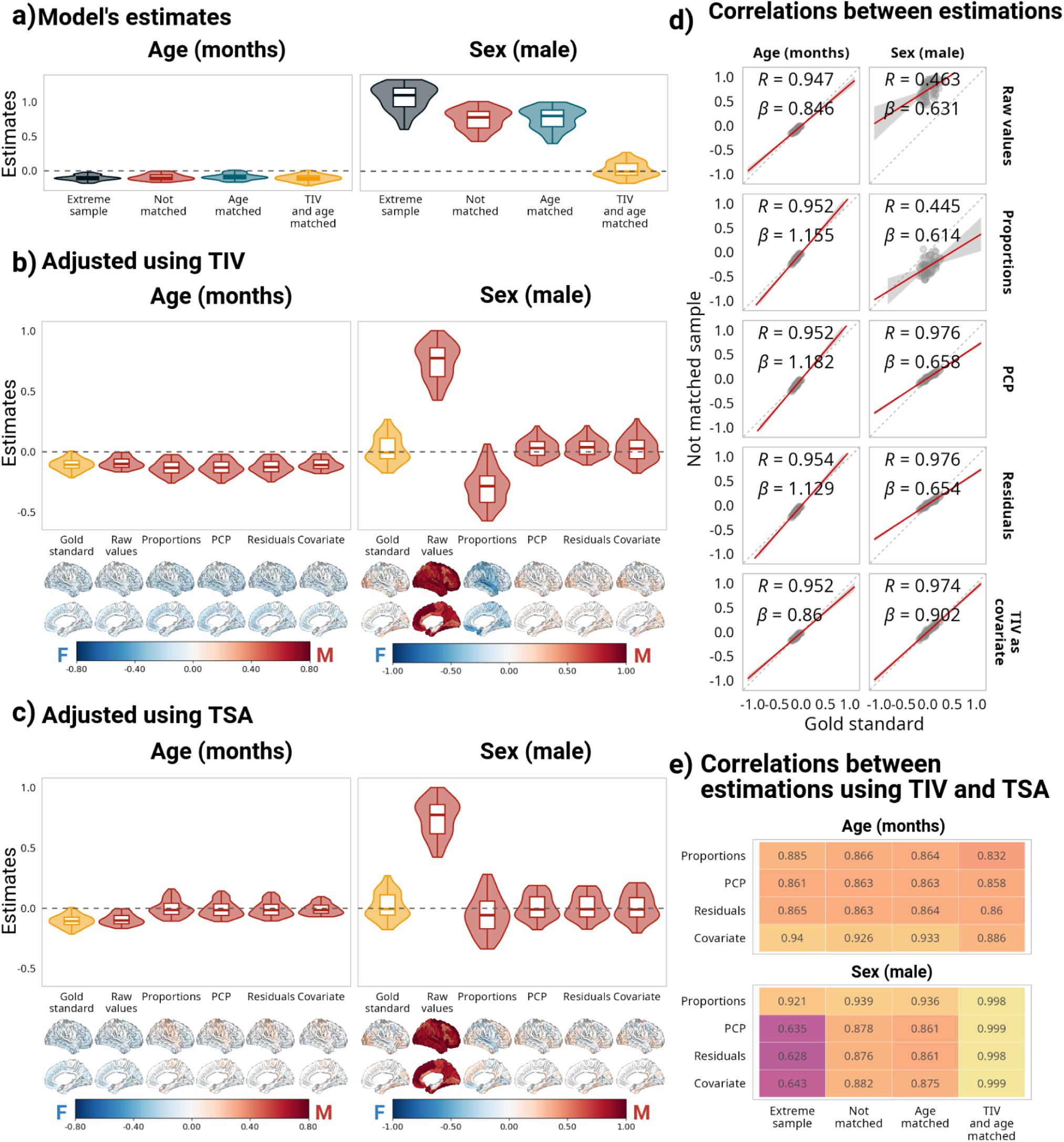
Impact of correction methods on model estimations based on surface area. **a**: Distribution of estimates for all samples. **b and c**: Comparison of estimates for the matched sample without adjustment and the estimates after using different adjustment methods for the not matched sample. **b** presents the estimates using TIV for the corrections, and **c** reflects the same results using TSA. Below the estimates are the corresponding cortical projections of the same values. **d**: Correlations between estimates for the matched sample without adjustment and the estimates after using different adjustment methods for the not matched sample. **e** Correlations between the estimates using different global measures for adjustment. For a comparison of all the estimates with the different global metrics, see Supplementary Figures. 5 and 6.

### Regional cortical thickness

Regional CT was negatively associated with age across all samples. Sex estimates in the age matched, not matched, and extreme samples leaned towards negative values, suggesting that CT is overall larger in females, while in the matched sample, this distribution was centered around zero (Figure 4a). Proportions was the only adjustment method that affected the age estimations. Both PCP and residuals method estimates were highly correlated with the gold standard, however, using TIV as a covariate yielded better results, without leading to a high degree of over or underestimations (Figure 4b). Note the regression slopes in Figure 4d for which TIV yielded I values close to one, indicating excellent correspondence to gold standard estimates. Using MCT as the adjustment metric led to a shift in the age estimates (Figure 4c left panel). Sex estimates were centered around zero for all methods, however, the correlations with the gold standard were below ρ = 0.7.

**Figure 4.**
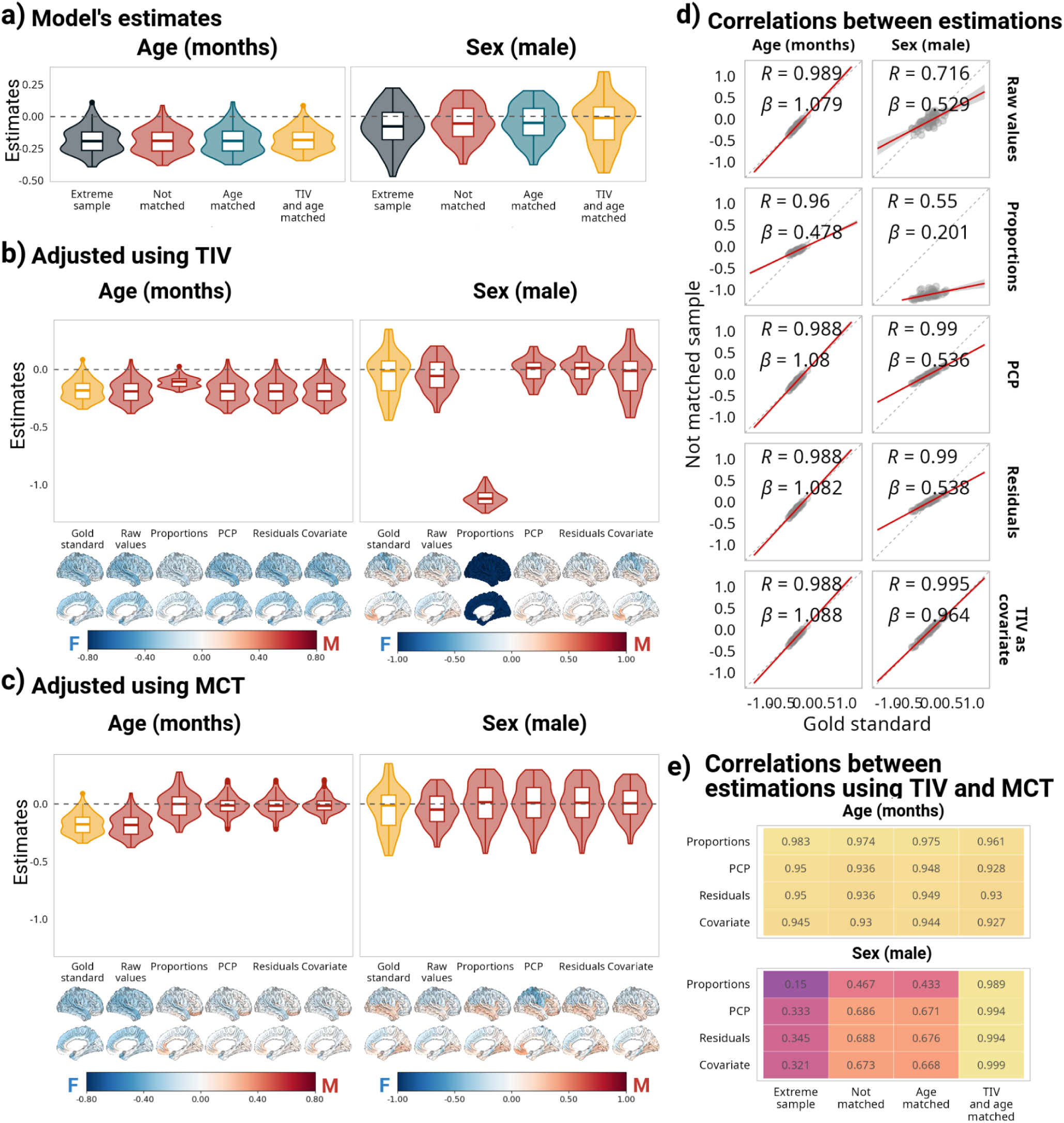
Impact of correction methods on model estimations based on cortical thickness. **a**: Distribution of estimates for all samples. **b and c**: Comparison of estimates for the matched sample without adjustment and the estimates after using different adjustment methods for the not matched sample. **b** presents the estimates using TIV for the corrections, and **c** reflects the same results using MCT. Below the estimates are the corresponding cortical projections of the same values. **d**: Correlations between estimates for the matched sample without adjustment and the estimates after using different adjustment methods for the not matched sample. **e** Correlations between the estimates using different global measures for adjustment. For a comparison of all the estimates with the different global metrics, and correlations between the gold standard and the different adjustment methods in the not matched sample, see Supplementary Figures 7 to 9.

### Vertexwise analyses

We extracted cortical metrics for 327,684 vertices. However, only 298,901(87.8%) of these were analyzed, including only vertices for which at least 80% of the participants in the full sample had non-missing values across the different smoothing levels.

### Vertexwise Surface Area

For 0 mm smoothing, SA estimates followed the same pattern as that of regional SA estimates (Figure 5a; full comparison in Supplementary Figure 10). None of the adjustment methods had a significant effect on the age estimations when TIV was used as the adjustment metric (Figure 5b), but they shifted from more negative values to being centered around zero when TSA was used for correction. Residuals, PCP, and adding TIV as a covariate centered the sex estimates around zero, mirroring the results obtained in the matched sample. Nonetheless, both residuals and PCP led to a decrease in the variability of the estimates, with the same biased underestimation as discussed before (Figure 5d). Across the cortex, the adjusted sex estimates correlated with gold standard estimates with ρ = 0.82 for PCP, and ρ = 0.87 for residuals and adding TIV as a covariate (Figure 5d). Interestingly, using TSA as the adjustment variable yielded very similar results (Supplementary Figure 11); however, in this case, the proportions method also shifted the sex estimates to be centered around zero. Despite this, it is worth noting that the correlations with the estimates obtained in the matched sample were lower (ρ = 0.57 for proportions, ρ = 0.75 PCP, ρ = 0.81 for residuals, and ρ = 0.82 for adding TSA as a covariate; see Supplementary Figure 12. Supplementary Figure 37 includes the correlations between the estimates obtained using TIV and TSA as the global metric).

**Figure 5.**
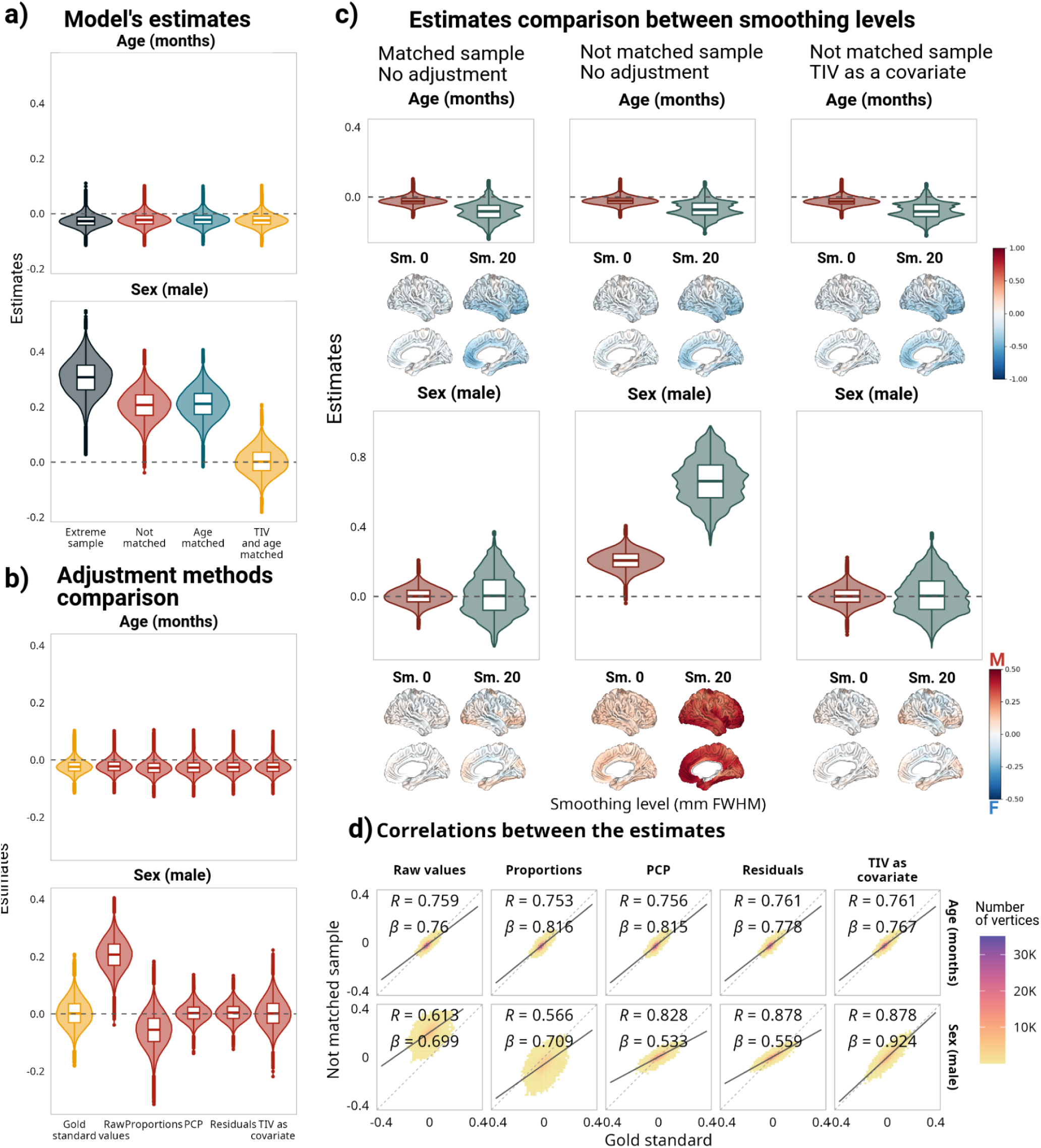
Impact of correction methods on model estimations based on vertexwise surface area with and without smoothing. **a**: Distribution of estimates for all samples. **b**: Comparison of estimates for the matched sample without adjustment and the estimates after using different adjustment methods for the not matched sample. **c:** comparison between the distribution of the estimates using data without smoothing versus data smoothed at 20 mm FWHM. Below each estimate are the corresponding cortical projections of the same values. **d**: Correlations between estimates for the matched sample without adjustment and the estimates after using different adjustment methods for the not matched sample. The analyses performed on data smoothed at 20 mm FWHM are included as Supplementary Figures 15 to 18.

For the impact of smoothing on the results, the data smoothed at 5-25 mm FWHM yielded similar results regarding the general observed patterns (Supplementary Figure 13 for adjustments with TIV, and 14 for adjustments with TSA). Here, we use the no smoothing (0 mm) and 20 mm FWHM smoothing levels to highlight the overall findings from this analysis (Figure 5c). We observed two remarkable differences: First, the magnitude of the estimates, and thus variance increased with increase in smoothing levels, which led to large biases in regional sex estimations. Secondly, when using a 20 mm FWHM smoothing level, the correlations between gold standard and not matched sample estimates increased to ρ > 0.95 for the age and sex estimates after applying different correction methods (except for the proportions method) when using TIV as the adjustment variable, and ρ > 0.9 when using TSA (Supplementary Figures 17 and 18). Note that the analyses performed on vertexwise cortical volumes yielded very similar results to those obtained for surface area, and are included as Supplementary Figures 19 to 26 for brevity.

### Vertexwise cortical thickness

For 0 mm FWHM smoothing, vertexwise CT estimates followed the same pattern across samples as those of regional CT. In contrast to the findings for SA, the general pattern was similar across samples, even for the sex coefficient (Figure 6a; full comparison in Supplementary Figure 27). Using TIV as the adjusting metric, applying different adjustment methods did not have a significant effect on the age estimations. However, using MCT for adjustment led to a shift from more negative values to values centered around zero (Supplementary Figure 28). The residuals and PCP methods centered the sex estimates around zero, mirroring the results obtained in the matched sample, and further led to a decrease in the variability of the estimates. Across the cortex, the adjusted sex estimates correlated with gold standard estimates with ρ = 0.94 for PCP, ρ = 0.95 for residuals, and ρ = 0.96 for adding TIV as a covariate (Figure 6d). Using MCT as the adjustment variable led to different results, with all the methods centering the sex estimations around zero.

**Figure 6.**
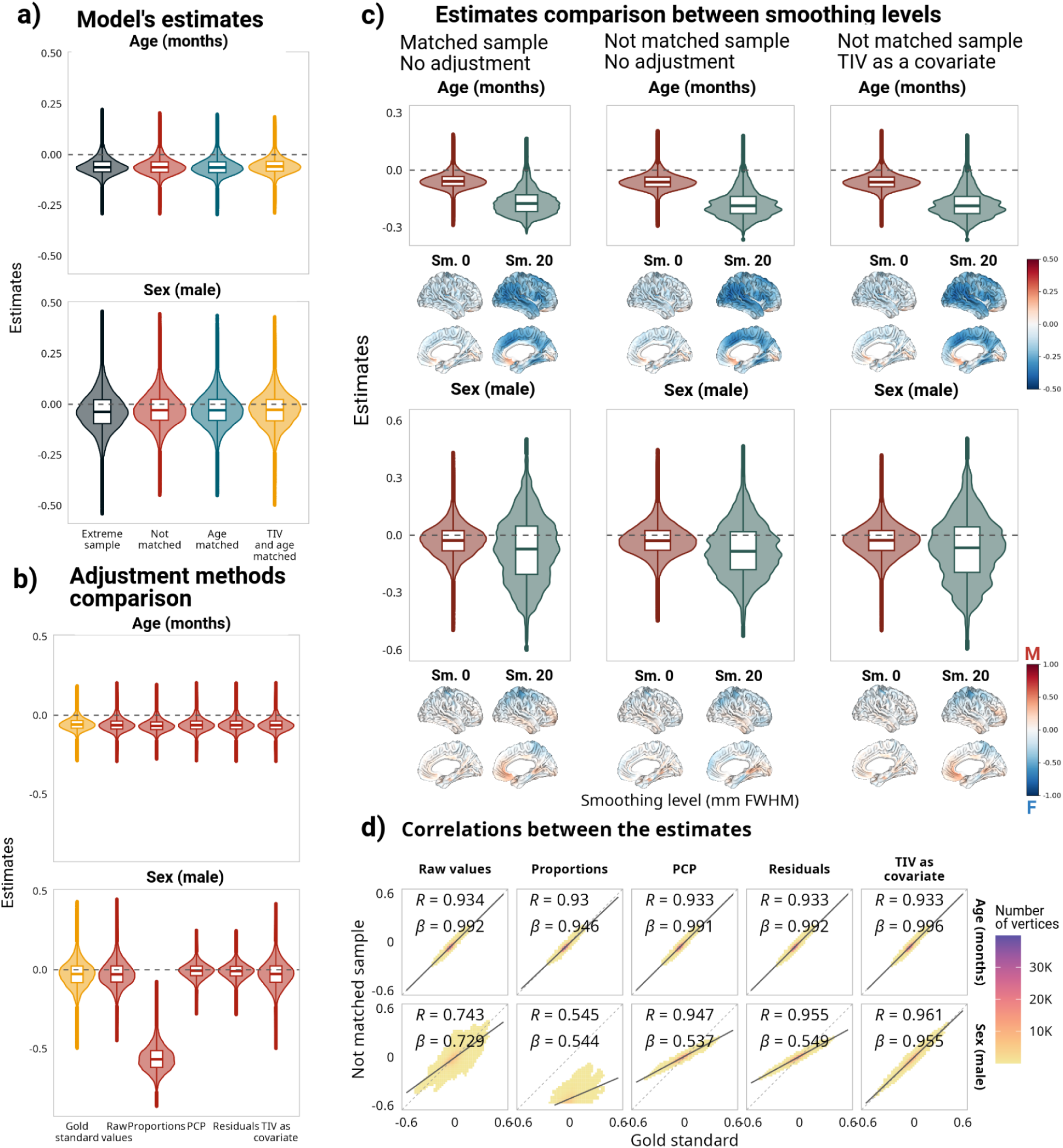
Impact of correction methods on model estimations based on vertexwise cortical thickness with and without smoothing. **a**: Distribution of estimates for all samples. **b**: Comparison of estimates for the matched sample without adjustment and the estimates after using different adjustment methods for the not matched sample. **c:** comparison between the distribution of the estimates using data without smoothing versus data smoothed at 20mm FWHM. Below each estimate are the corresponding cortical projections of the same values. **d**: Correlations between estimates for the matched sample without adjustment and the estimates after using different adjustment methods for the not matched sample. For additional information on the analyses performed with data smoothed at 20mm FWHM and using MCT, see Supplementary Figures 27 to 36.

For the impact of smoothing on the results, the data smoothed at 5-25 mm FWHM revealed that increase in smoothing leads to i) an increase in the variability of the estimates, and ii) the age and sex estimates leaning towards more negative values in both matched and non matched samples (Supplementary Figure 35). Contrary to the smoothing dependent bias in sex estimations in SA, applying different adjustment methods to the smoothed data resulted in the same trends previously observed in non-smoothed data across smoothing levels indicating minimal impact of smoothing on CT results after correction.

### Voxelwise DBM

For voxelwise DBM values, instead of only focusing on the gray matter (GM), we analyzed the whole brain using BISON^78^ segmentation, which provided further insight in both GM and white matter (WM). In both GM and WM, the age estimates leaned towards negative values across all samples. Cortical GM sex estimates (Supplementary Figure 38) were highly correlated between the matched and the non matched sample (⍴ = 0.73; Figure 7); however, most estimates were over- or underestimated in the not matched sample (note the regression slopes presented in Figure 7). A similar trend was present in the deep GM (Supplementary Figure 39), while in this case, the correlation was ⍴ = 0.48 and the estimates were mostly underestimated in the non matched sample in comparison to the gold standard. WM estimates of the two samples were correlated at ⍴ = 0.51 (Figure 7), and were either over- or underestimated in the non matched sample (Supplementary Figure 40). Interestingly, in the case of the ventricles (i.e. areas that can be indicative of atrophy), there were no correlations between the sex estimates obtained for the two samples (Figure 7).

**Figure 7.**
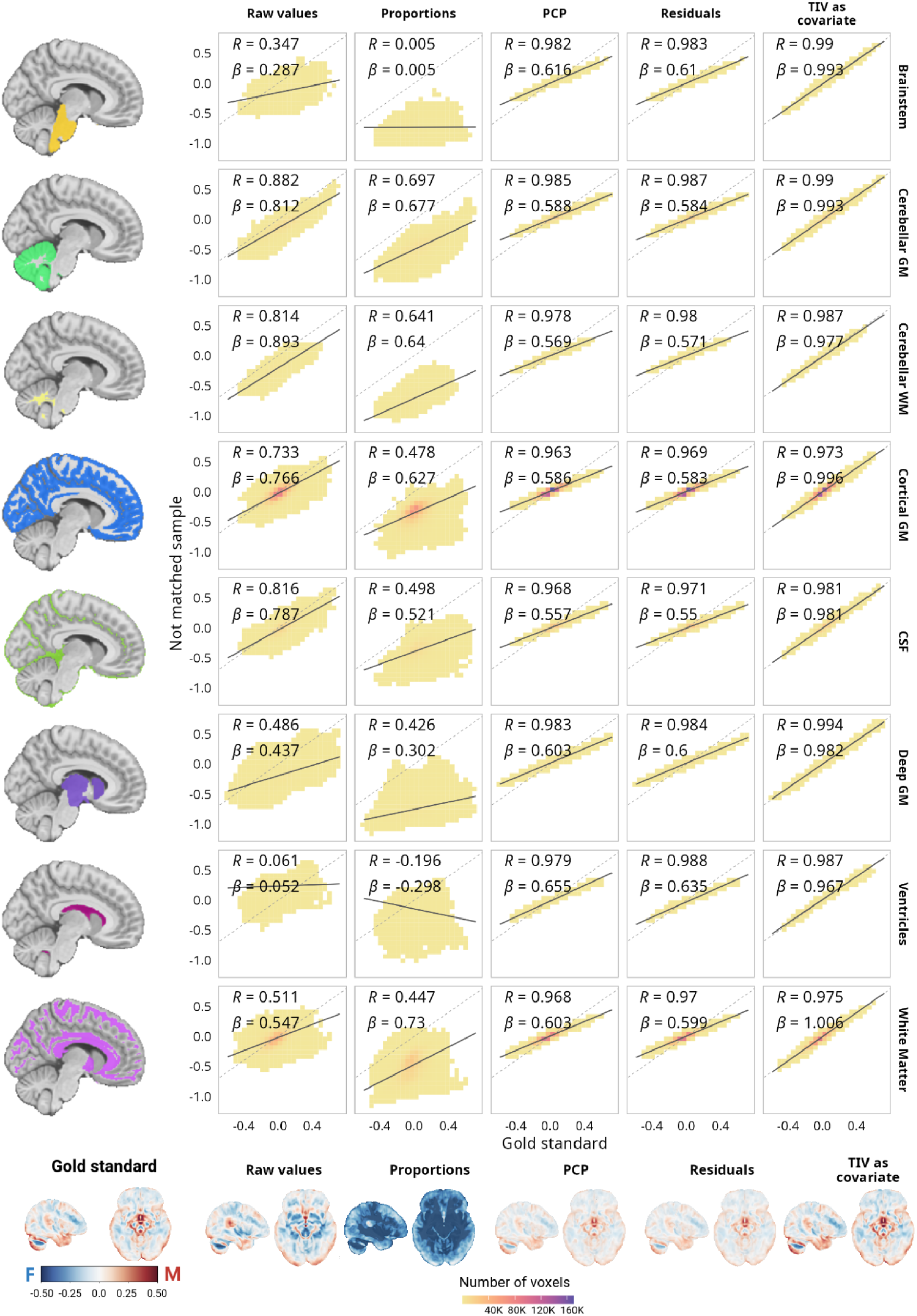
Correlations between the sex estimates for the matched sample without adjustment and the estimates after using different adjustment methods for the not matched sample using voxelwise Deformation Based Morphometry (DBM) values across different tissue types. All the brain projections are in the same colorscale. The same correlations for the age estimates are presented in Supplementary Figure 41. For a full comparison between the samples and methods in gray and white matter, see Supplementary Figures 38 to 40.

The different correction methods behaved similarly at voxel level across different tissues. First of all, the adjustments did not have a significant impact on the age estimations (Supplementary Figure 41). In the case of the sex estimates, the proportions method not only reversed the direction of the estimations but also inflated them. Both residuals and PCP methods yielded very similar results, with ⍴ > .96 across all tissues. However, they led to a decrease in variability across the estimates and the values were still over or underestimated. Adding TIV as a covariate generated the optimal results, with a correlation between the non matched sample and the gold standard of ⍴ > .97 across all the tissue types, as well as almost no over or underestimation bias.

### Biases in the estimations

Finally, we assessed the bias on the sex estimations that remained after using each correction method (using TIV as the adjusting variable), both in terms of magnitude and spatial distribution. Figure 8 shows the results at regional, vertexwise, and voxelwise analysis levels. Overall, the spatial distribution of the biases was similar across the methods. However, in terms of magnitude, adding TIV as a covariate was best at removing the biases from estimates across the brain. Residuals and PCP results were virtually identical and both left higher biases after correction compared to the TIV adjustments while following a similar pattern to it. Interestingly, at both regional and vertex levels, biases in corrected estimates were larger for cortical thickness and smaller for surface area, suggesting a more linear scaling in case of surface area across the regions. For the volumetric and cortical thickness data, the biases were mainly localized on dorsal areas, while for regional DBM, we observed a positive bias (undercorrection, i.e., a higher bias towards male estimates than suggested by gold standard) on temporal regions when using the residuals or PCP methods. For volumetric and DBM data, overall, the bias was stronger in subcortical regions. It is worth noting that for vertexwise data, higher smoothing levels were associated with increased biases.

**Figure 8.**
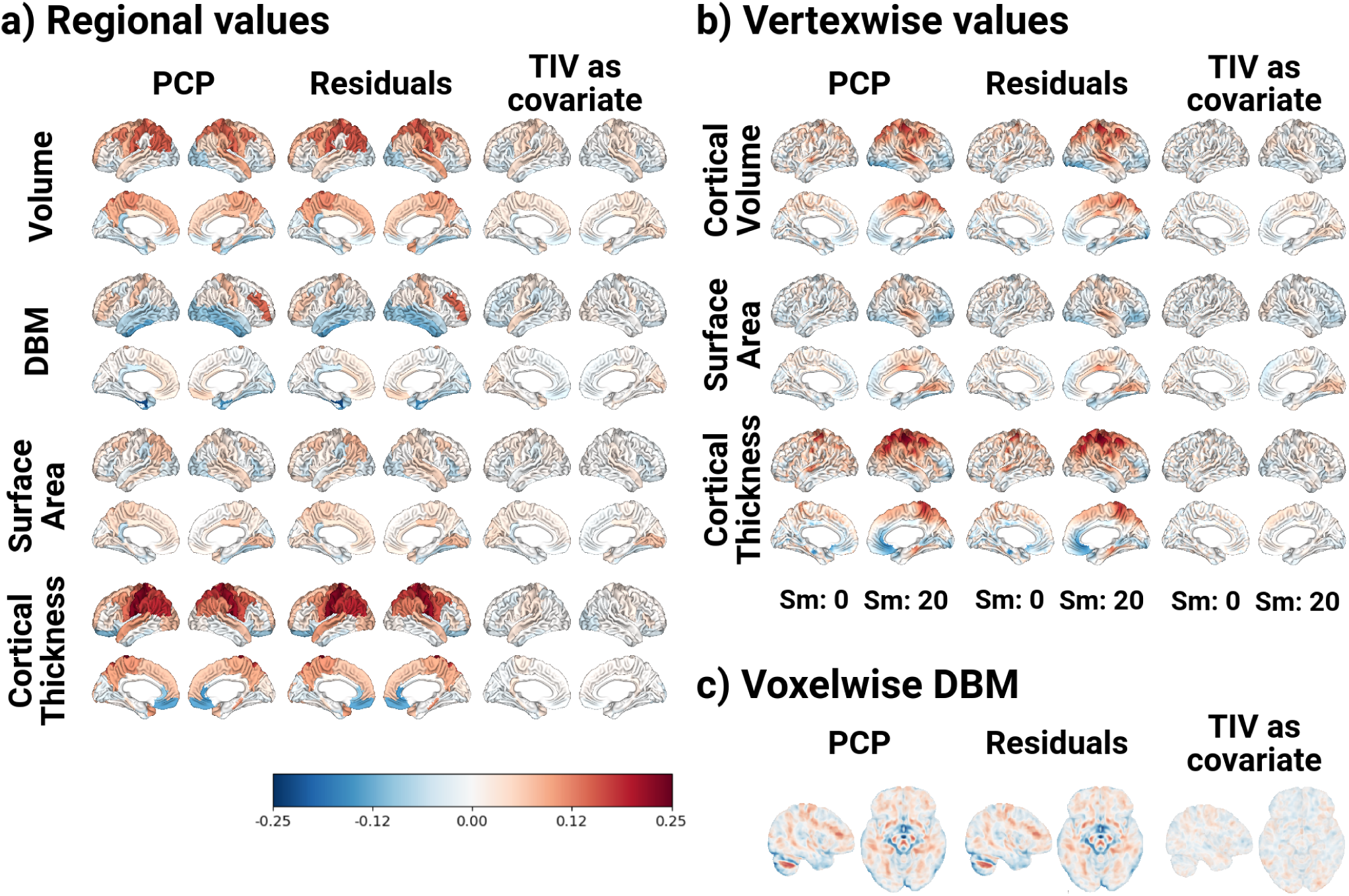
Residual biases after the implementation of the different correction methods. a) biases in the regional estimations. b) biases on the vertexwise estimations. For each correction method, the left hemisphere (on the left) represents the biases when the data was not smoothed, and on the right hemisphere, when the data was smoothed 20mm FWHM. c) biases at the voxel level based on deformation based morphometry data.

## Discussion

In this study, we evaluated the performance of different methods in adjusting MRI-derived brain structural measures to control for the effect of brain size in neuroimaging studies with a focus on age and sex estimation in later adulthood (45-80 years olds). Taking advantage of the UKBB dataset, we implemented a matching approach to create a subsample of participants where females and males were perfectly matched by age and TIV, thus having a **gold standard** to compare four different adjustment methods, namely the proportions, residuals, power corrected proportions, and adding TIV as a covariate. Each method was evaluated at regional, vertex, and voxel levels, using a consistent linear regression approach for all analyses and comparing the standardized beta coefficients between methods and samples.

Our results suggest that, regardless of the metric of interest, adding TIV as a covariate in a regression model (per region/vertex/voxel) is the best method for removing the effect of TIV on sex differences in aging trajectory models, compared to a gold standard where we enforced the same TIV distribution for both sexes. Importantly, in addition to TIV, we also evaluated using total SA (TSA), for adjusting regional and vertexwise SA, mean CT (MCT) for adjusting CT measures, and total cortical volume for vertexwise volume. In all cases, these metrics impacted the age estimations, suggesting that using these measures for adjustment biases study findings and the results will be suboptimal at best and biased at worst in comparison to TIV based adjustment. This highlights the importance of using a measure that is consistent across the aging process, instead of one that is also affected by it ^19,73^.

Previous studies had evaluated various adjustment methods for volumetric measures in different contexts ^23,25,26,26–34^. The most common comparisons were between proportions and residuals methods, and in some cases, the covariate method. While these studies were mainly based on relatively small sample sizes (under 100 participants), overall, it was recognized that, apart from very specific cases where the goal of the study was to examine differences in the proportion of volume occupied by a certain structure, residuals and covariate methods yielded better results. Most of these studies focused either on a diseased versus control population, or only on sex differences; in this study, we also highlighted how each method affects age estimations in the context of aging studies. This method is recommended due to its greater flexibility when it is necessary to incorporate additional covariates^23,30^. Importantly, using TIV as a covariate has the advantage that, while removing the effect of brain size from other estimations, it also allows us to study how TIV might directly impact the phenomenon of interest with biological significance rather than being a covariate of no interest with indirect relationship.

To our knowledge, this study is the first attempt to perform a systematic analysis on the effects that different adjustment methods can exert on estimating sex differences using different MRI-derived brain measurements; i.e. cortical thickness, surface area, volumes, and DBM compared to gold standard matching method. The effects we observed with surface area were very similar to those observed on volumetric data, which is expected as surface area and brain volume are closely related measures ^10,81^ . Cortical thickness results were particularly interesting, where sex estimate correlations between the uncorrected non-matched and gold standard samples were higher than those of the other metrics, and the impact of using different correction methods was less evident. A potential explanation is that CT is poorly correlated with body and head size measures^10,82–85^. This is also the first systematic investigation of the effect of brain size adjustments on DBM. DBM processing includes a linear registration step which linearly scales the brain to the stereotaxic space prior to estimating voxelwise atrophy. As seen in the results, this scaling is not equivalent to adjusting for TIV and additional adjustment is necessary.

The various methods evaluated in this study take different elements into consideration. For instance, the proportions method was originally designed to examine the percentage of intracranial volume occupied by a given brain region. Meanwhile, the residuals method was intended for group comparisons in the context of a diseased versus a control population ^40,41^, which would potentially hinder its use if the goal of a study is not contrasting different populations. The PCP method is based on considering the non-linear scaling relationship between a brain region and a global size measure^42^ (i.e. brain allometry ^36,38^). Importantly, in our previous work, we reported that brain allometry is consistent between sexes and across a wide spectrum of brain sizes^19^. Meanwhile, the covariate method linearly residualizes any metric of interest by the adjustment metric. Interestingly, this suggests that the introduced biases for smaller and larger TIVs are mainly linear regardless of allometry, and this linear relationship can vary between regions. In other words, within the age range under study, a linear relationship is the best representation of the scaling of the brain while neurodevelopmental stages warrant further investigations.

Evaluating the impact of various smoothing levels on vertexwise data, we found that independent of the selected adjustment method, an increase in smoothing level is associated with larger estimates both for age and sex, which leads to an increase in estimates’ variability and biases, in the three analysed metrics. This is potentially due to the increased signal-to-noise ratio and disappearance of individual variability that result from smoothing, which increases the probability of obtaining significant group differences ^13^ and hence introducing less reliable local estimates in favor of capturing overall differences. While there is no straightforward answer to what an ideal smoothing level is^16^, it is advisable to consider the implications of using a given smoothing level for any analysis and considering lower smoothing levels when subtle regional variabilities are of importance to the question of interest.

We also acknowledge the limitations of our study. Despite being a large dataset for neuroimaging studies, the UKBB sample might not be fully representative of the general population, even within the area where it was collected ^20^. For instance, it has been noted that compared to the general population, the participants in the UKBB report lower smoking rates and alcohol intake, have fewer self-reported health conditions, are less likely to be obese, and more likely to live in less socioeconomically deprived areas ^57^. Additionally, from the population who participated in the imaging acquisition protocol, over 96% were Caucasian, which limits the generalizability of the findings to other ethnic groups.

Our study has several strengths that support the reliability of our findings. First, we had access to the UK Biobank dataset with over 35,000 imaging data points of participants ranging from 45 to 80 years old, which allowed us to create perfectly matched subsamples with over 10,000 participants in each subsample, thus assuring statistical power and robustness of our results. Second, we consistently used the same analytical methodology (i.e. linear regression) among different adjustment methods and metrics tested, assuring the comparability of our results. Third, visual quality control was part of our image preprocessing pipeline, ensuring the accuracy of the TIV estimations. Furthermore, other potential sources of error, such as excessive motion in the scanner, incidental findings, or incomplete field of view, were also excluded from our analyses.

In conclusion, performing a comprehensive systematic evaluation of four different TIV adjustment methods to assess sex differences in various neuroimaging metrics, we found that adding TIV as a covariate is optimal for removing the effect of brain size from sex differences estimations, at region, vertex, and voxel levels. This effect is more prominent in volumetric, surface area, and DBM calculations than for cortical thickness.

## Data Availability

All the data for this study comes from the UK Biobank dataset. The UK Biobank dataset is open access and can be requested from https://www.ukbiobank.ac.uk/use-our-data/apply-for-access/

## Authors contributions

**A.B.R.**: Conceptualization, Formal analysis, Interpretation of findings, Methodology, Visualization, Writing - original draft, Writing - review & editing. **R.M.**: Formal analysis, review & editing. **Y.Z.**: Conceptualization, Formal analysis, Interpretation of findings, Methodology, Visualization, Writing - original draft, Writing - review & editing. **M.D.**: Conceptualization, Formal analysis, Interpretation of findings, Methodology, Visualization, Writing - original draft, Writing - review & editing.

## Acknowledges

This research was conducted using the UKBB Resource under approved application 45551. We thank the UKBB participants and team for their work in collecting, processing, and disseminating these data for analysis. The authors also acknowledge use of Compute Canada (https://alliancecan.ca/en) resources for performing the image processing analyses in the presented work.

