## Supplemental figures 1-41 for "Brain Size: To Adjust or Not Adjust? It’s Not a Matter of If, but How"

### Supplementary Material

Brzezinski-Rittner, et.al.

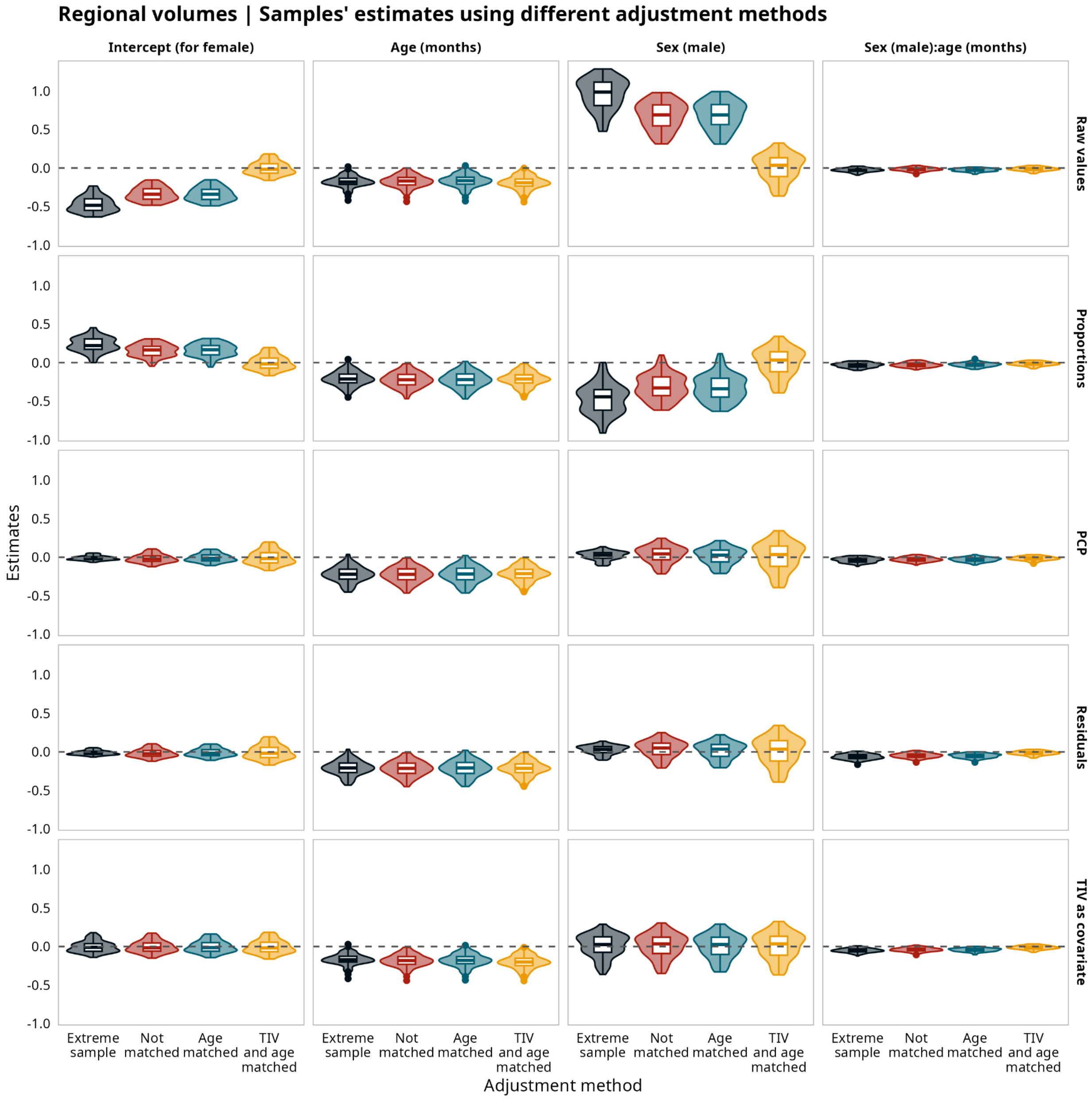


**Supplementary Figure 1.** Regional volumes. Model's estimates for regional volumes for all the samples and all the adjustment methods.

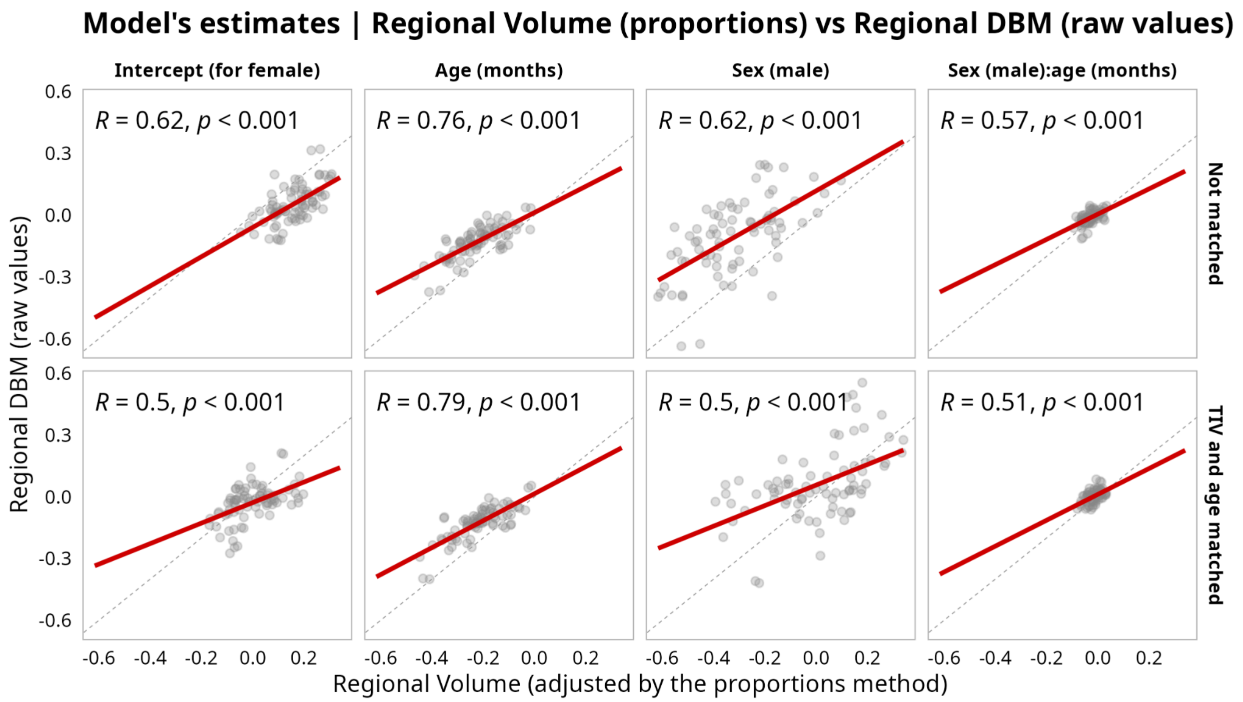


**Supplementary Figure 2.** Comparison between the model's estimates for volumetric data adjusted using the proportions method versus the estimates for regional deformation based morphometry data without adjustment.

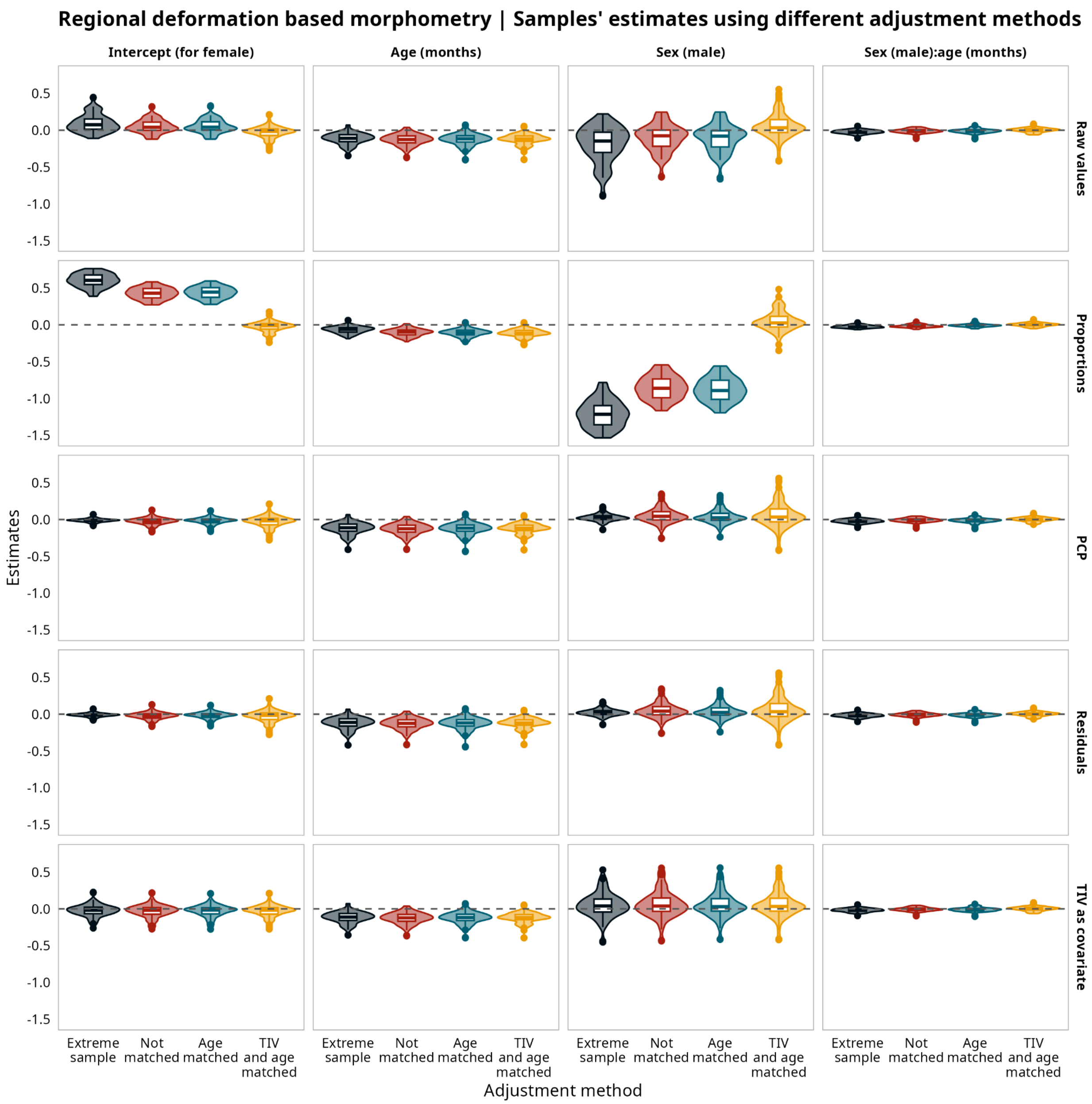


**Supplementary Figure 3.** Regional deformation based morphometry (DBM). Model's estimates for regional DBM for all the samples and all the adjustment methods.

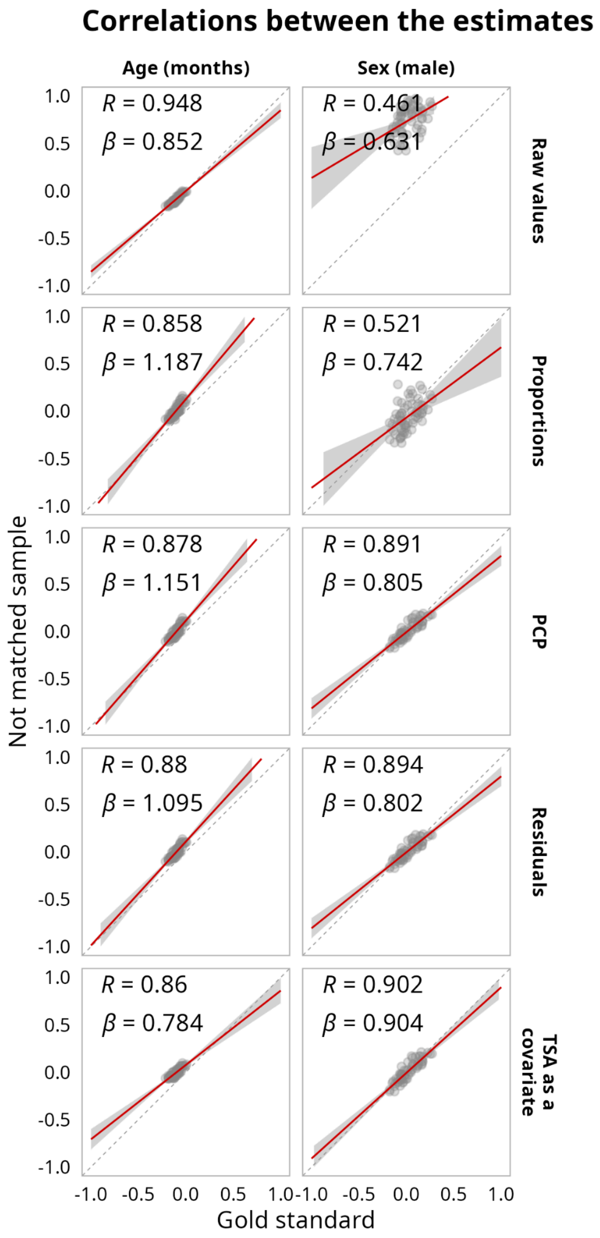


**Supplementary Figure 4.** Regional Surface Area. Correlations between the model's estimates for regional surface area for the matched sample without adjustment and the estimates after using different adjustment methods for the not matched sample using total surface area (TSA) as the adjustment metric.

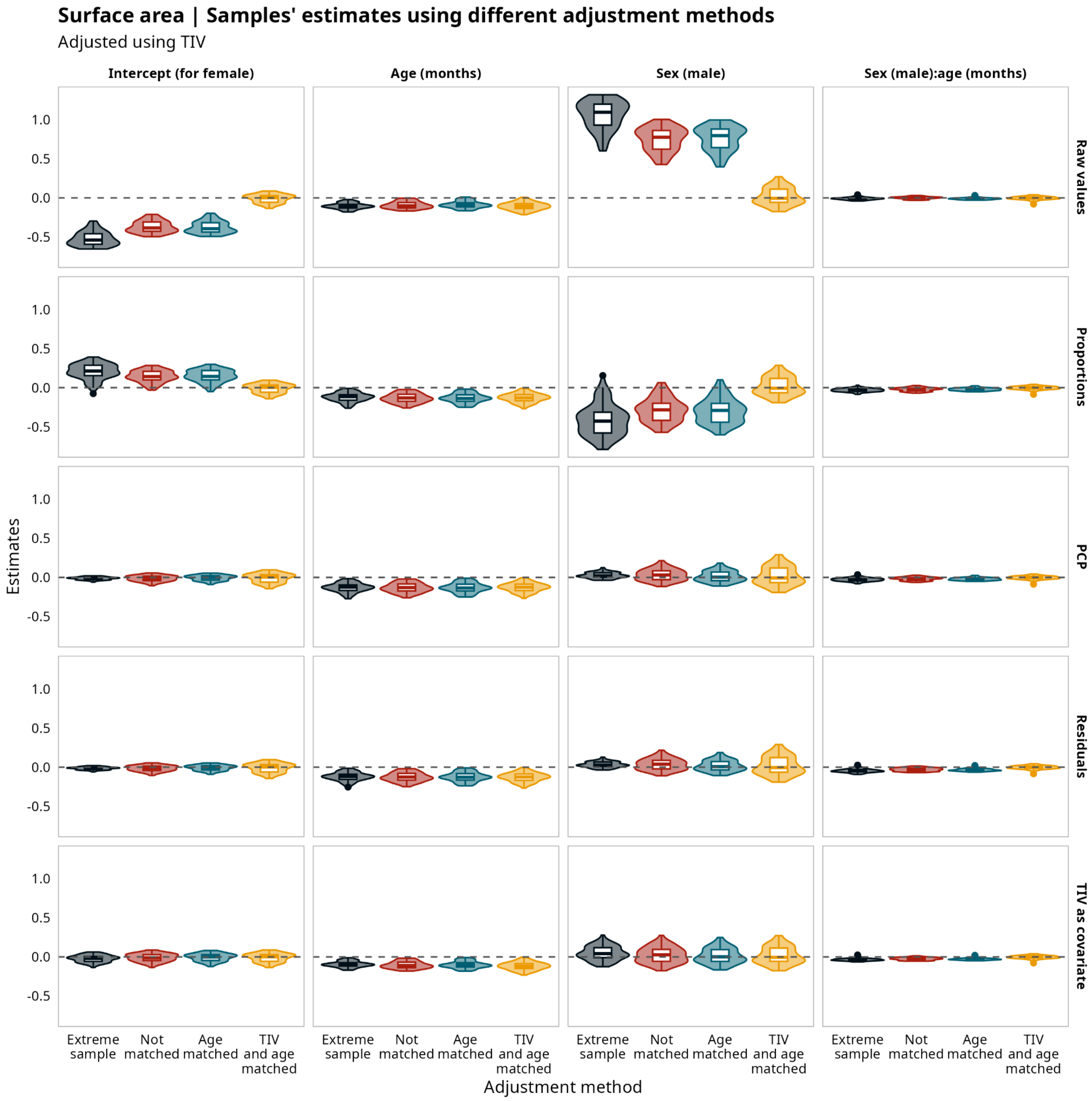


**Supplementary Figure 5.** Regional surface area. Model's estimates for regional surface area adjusted using total intracranial volume (TIV) for all the samples and all the adjustment methods.

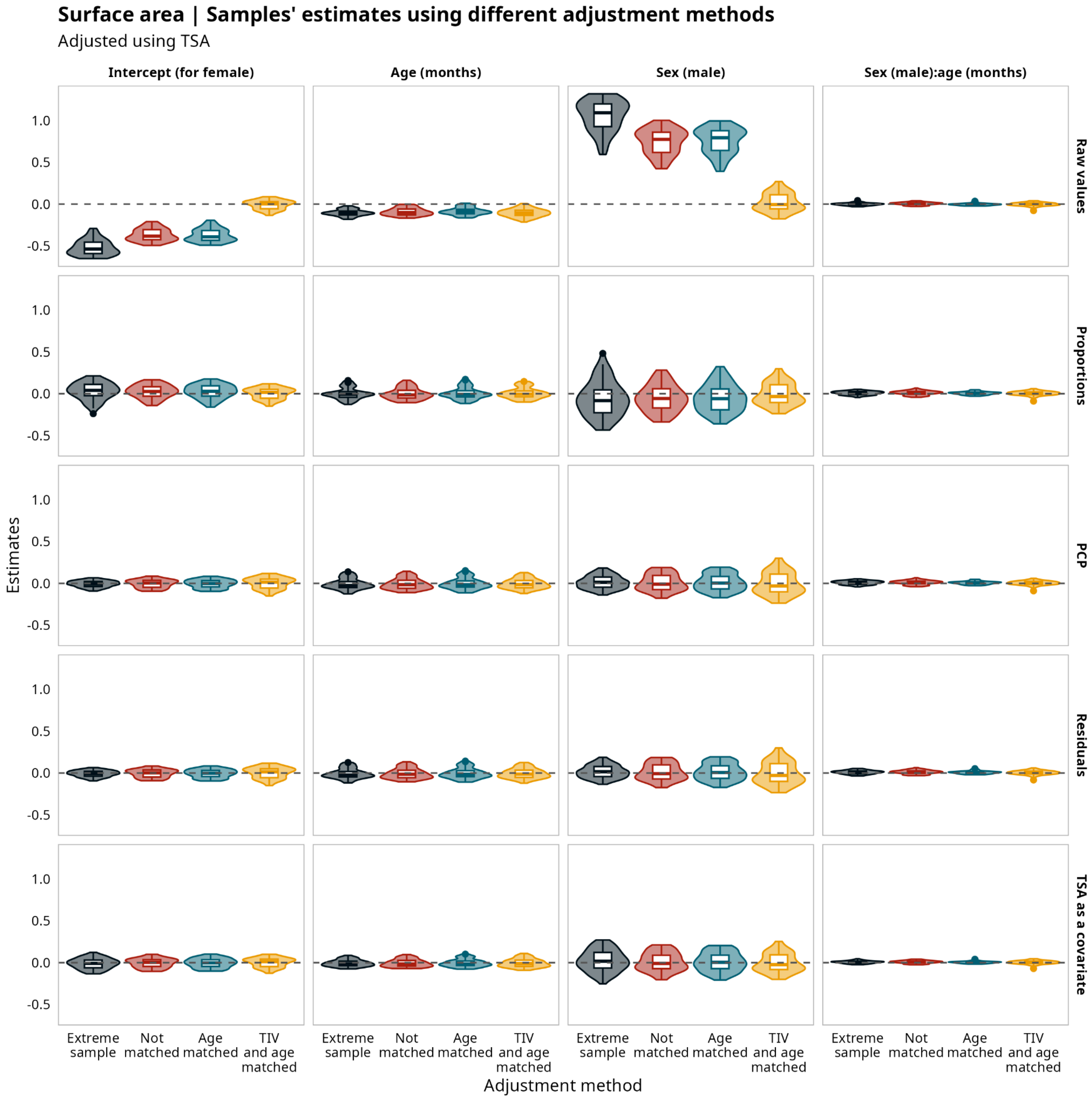


**Supplementary Figure 6.** Regional Surface Area. Model's estimates for regional surface area corrected using total surface area (TSA) for all the samples and all the adjustment methods.

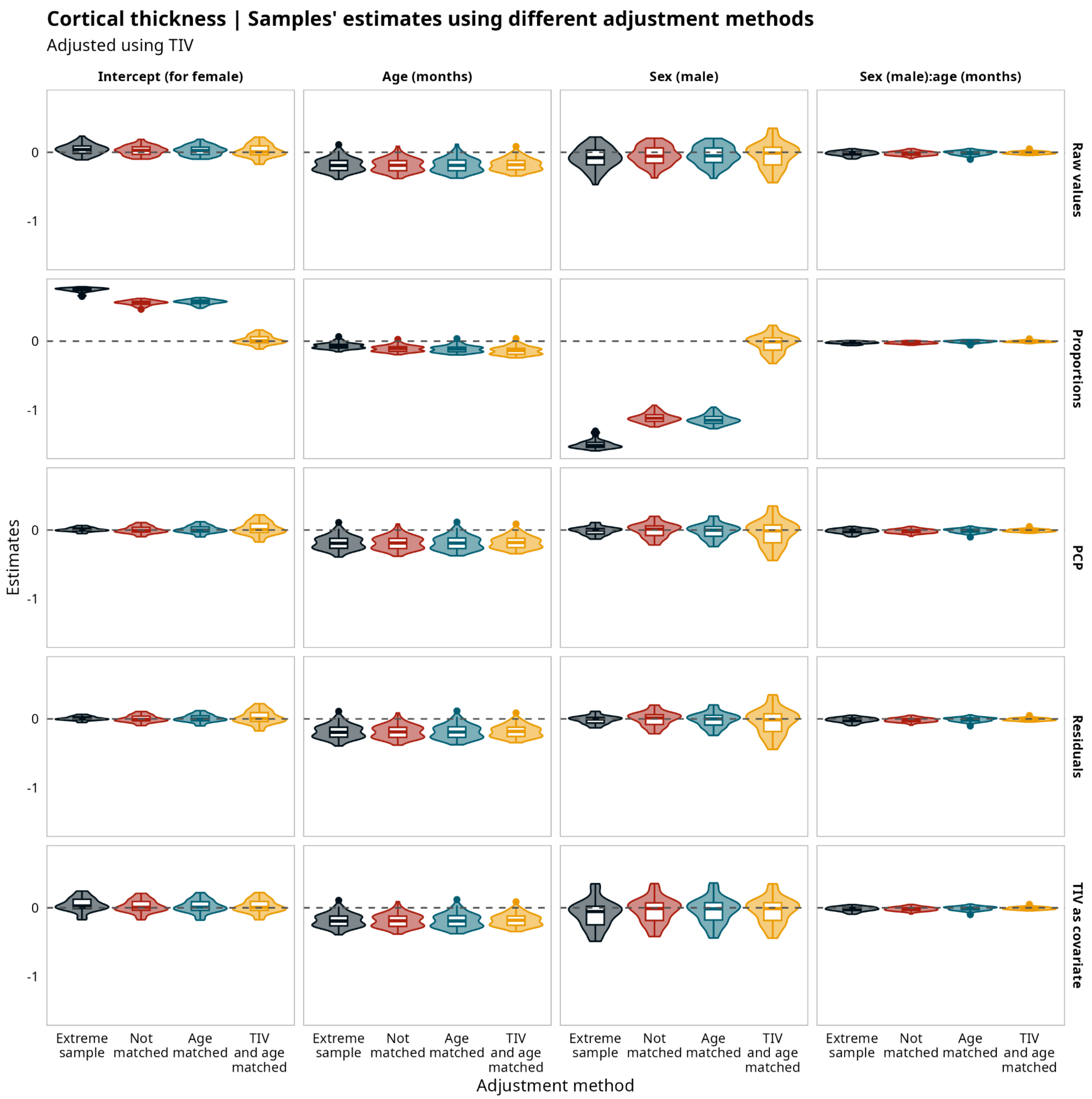


**Supplementary Figure 7.** Regional Cortical Thickness. Model's estimates for regional cortical thickness adjusted using total intracranial volume (TIV) for all the samples and all the adjustment methods.

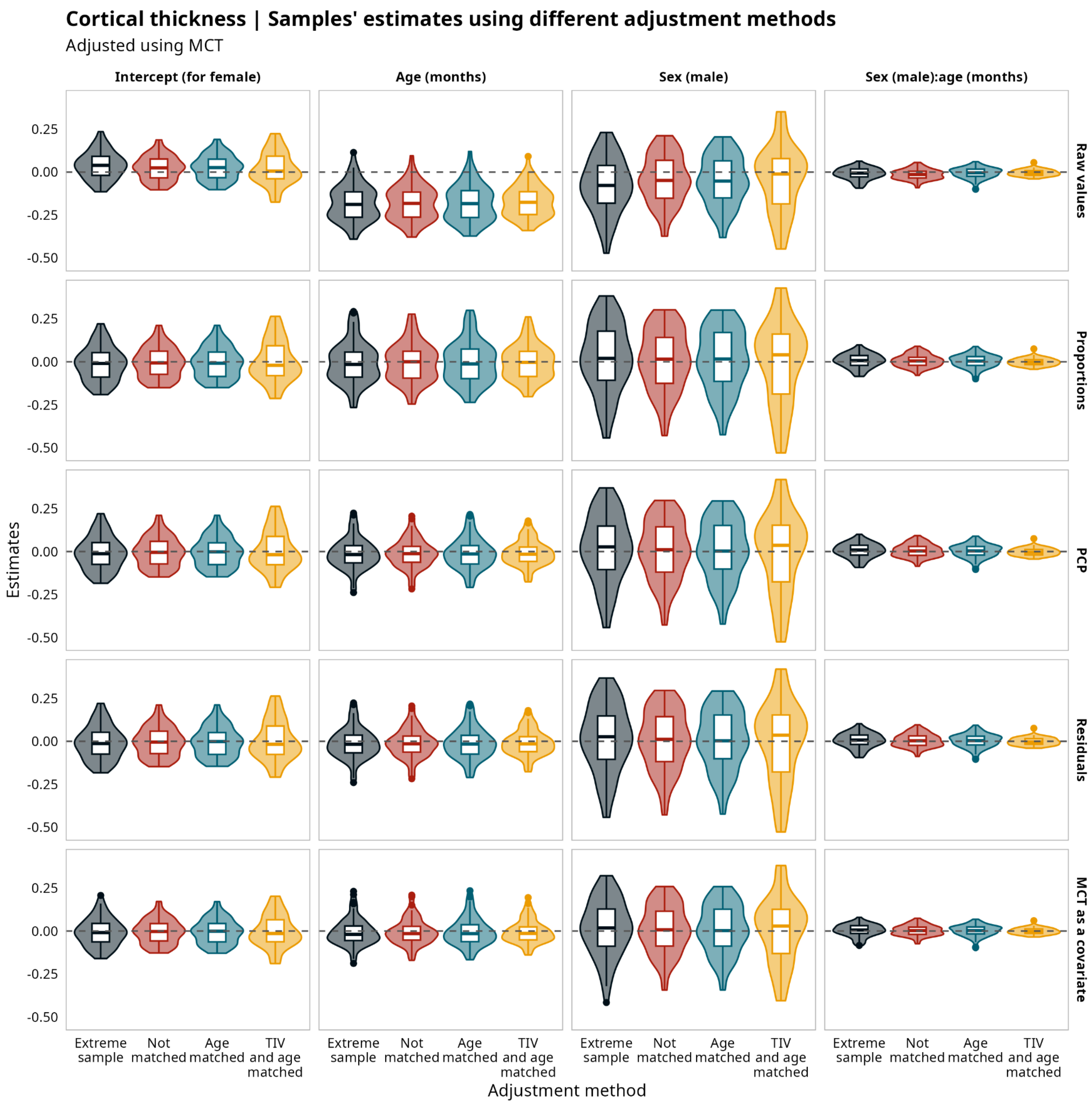


**Supplementary Figure 8.** Regional Cortical Thickness. Model's estimates for regional cortical thickness adjusted using mean cortical thickness (MCT) for all the samples and all the adjustment methods.

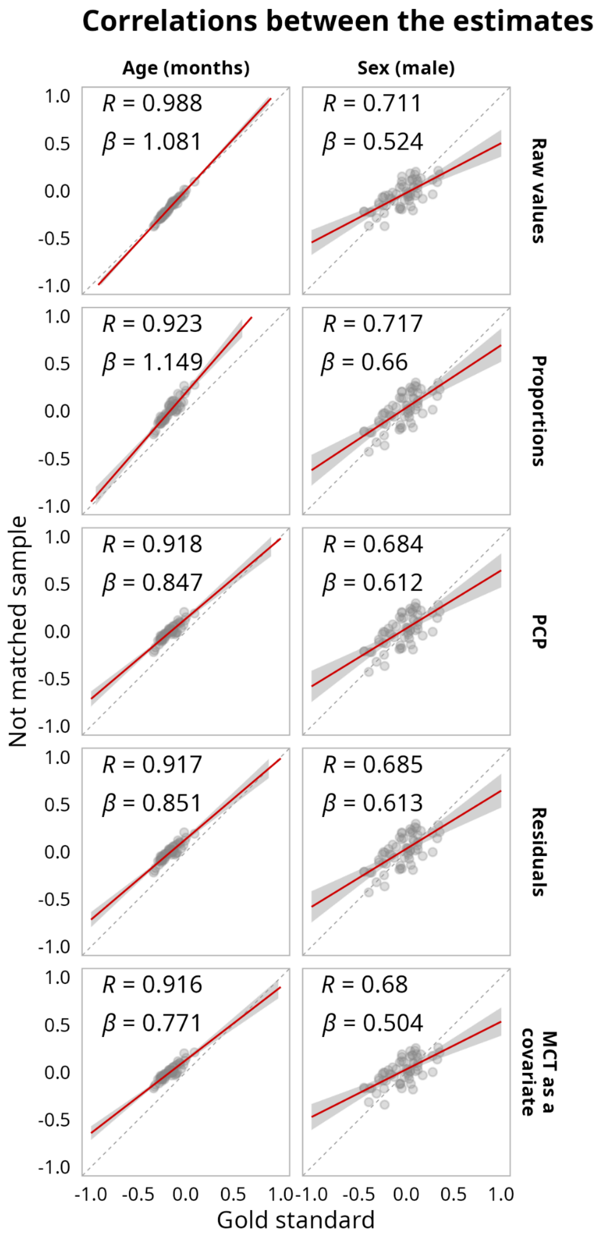


**Supplementary Figure 9.** Regional Cortical Thickness. Correlations between model's estimates for regional cortical thickness for the matched sample without adjustment and the estimates after using different adjustment methods for the not matched sample using mean cortical thickness (MCT) as the adjustment metric.

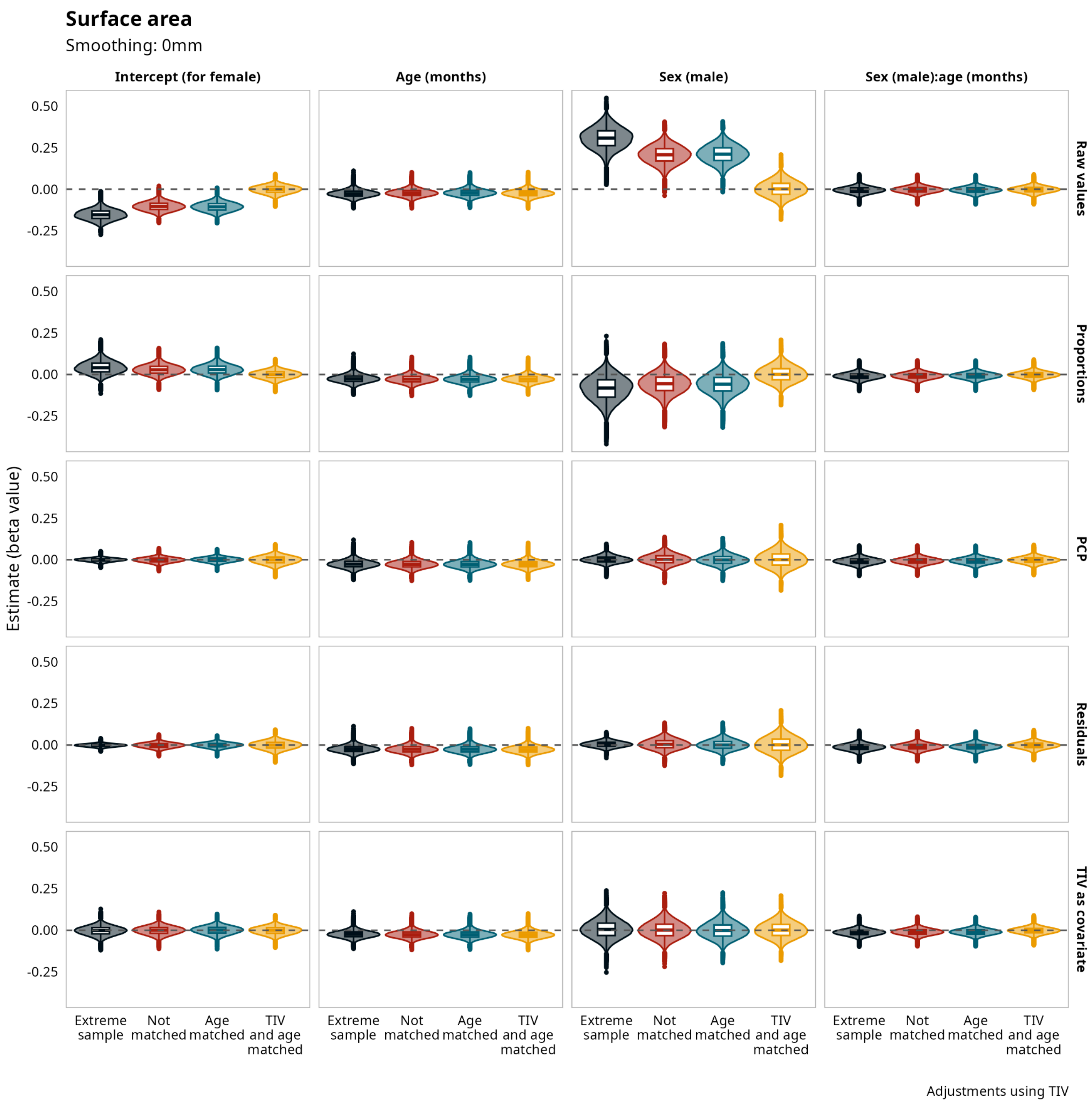


**Supplementary Figure 10.** Vertexwise Surface Area. Model's estimates for vertexwise surface area without smoothing corrected using total intracranial volume (TIV) for all the samples and all the adjustment methods.

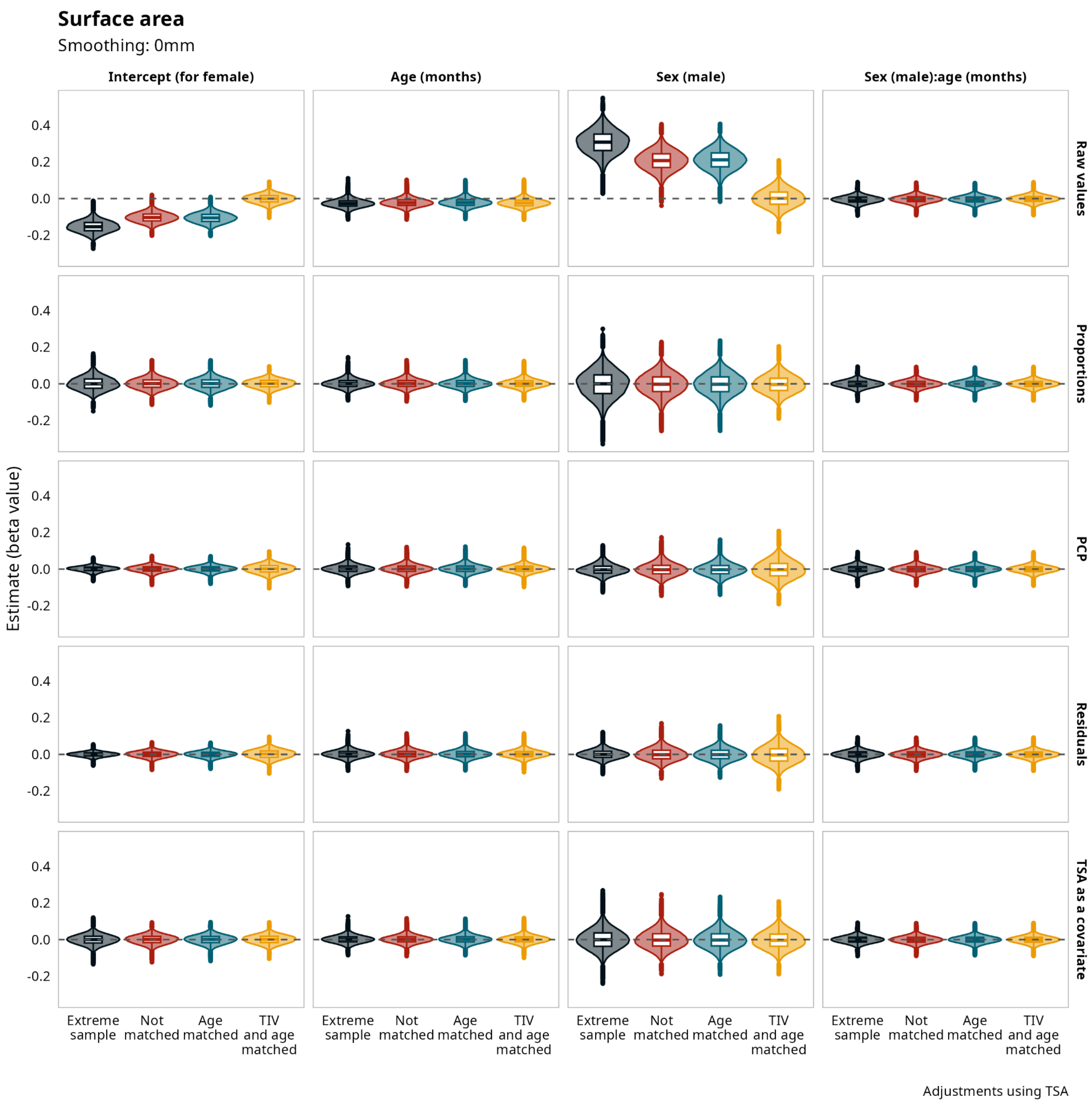


**Supplementary Figure 11.** Vertexwise Surface Area. Model's estimates for vertexwise surface area without smoothing corrected using total surface area (TSA) for all the samples and all the adjustment methods.

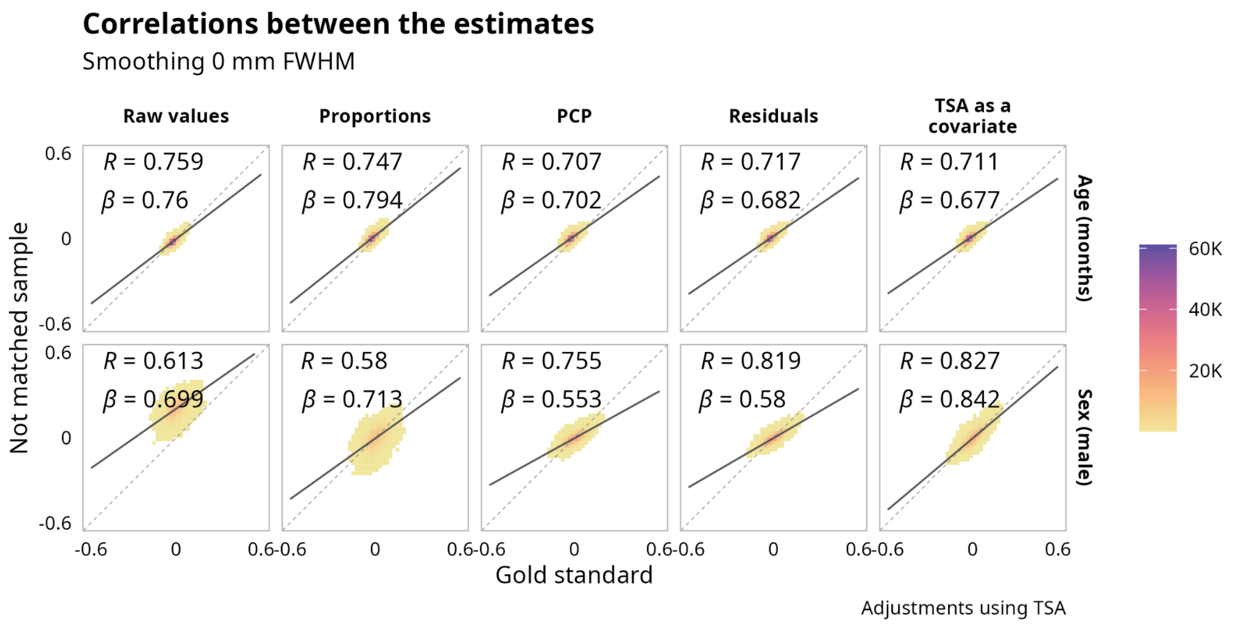


**Supplementary Figure 12.** Vertexwise Surface Area. Correlations between estimates for the matched sample without adjustment and the estimates after using different adjustment methods with total surface area (TSA) and the not matched sample, with data without smoothing.

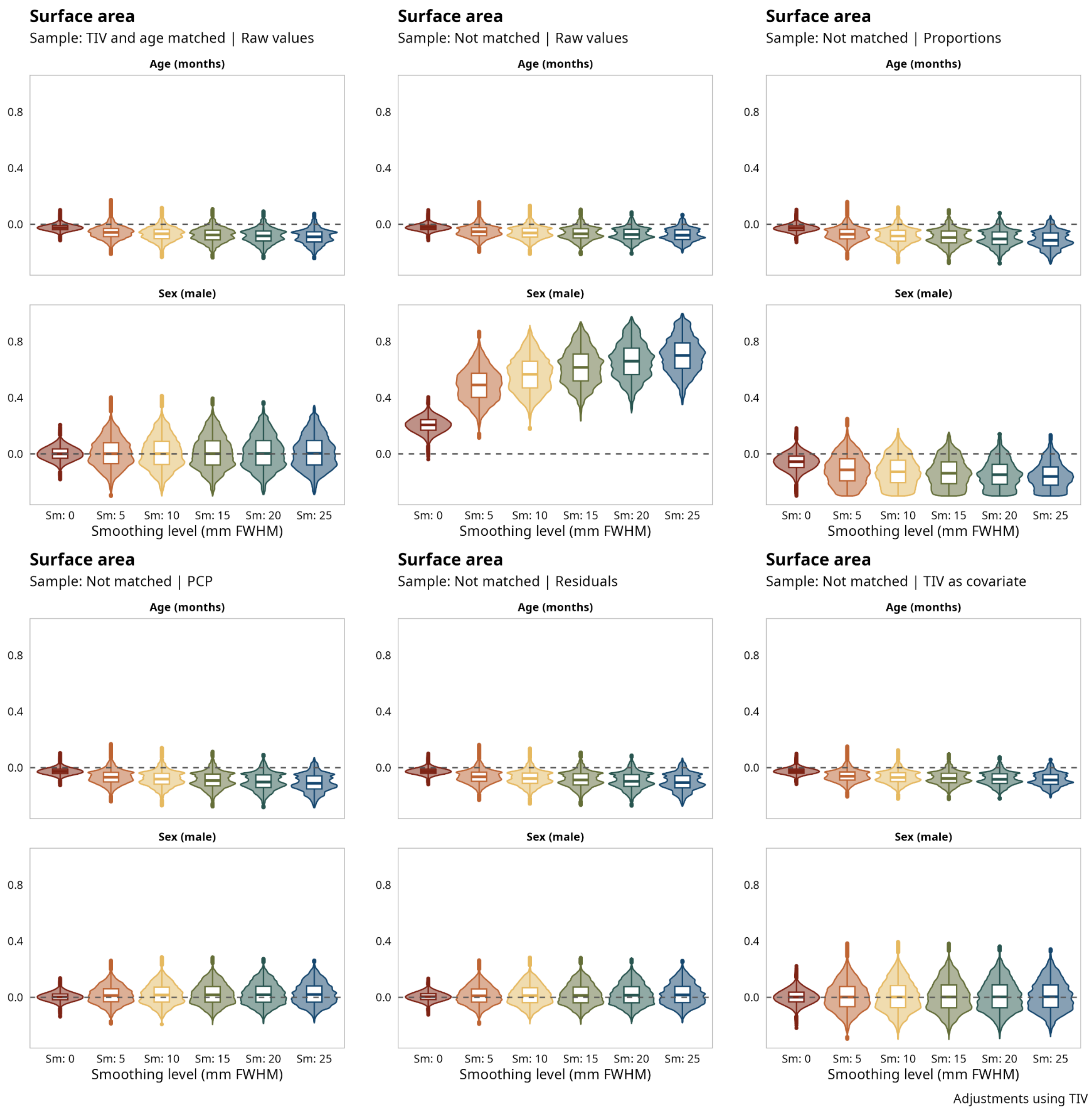


**Supplementary Figure 13.** Vertexwise Surface Area. Effect of different levels of smoothing on age and sex estimation, using the raw values in the mached and not matched samples and adjusting using total intracranial volume (TIV) in the not matched sample.

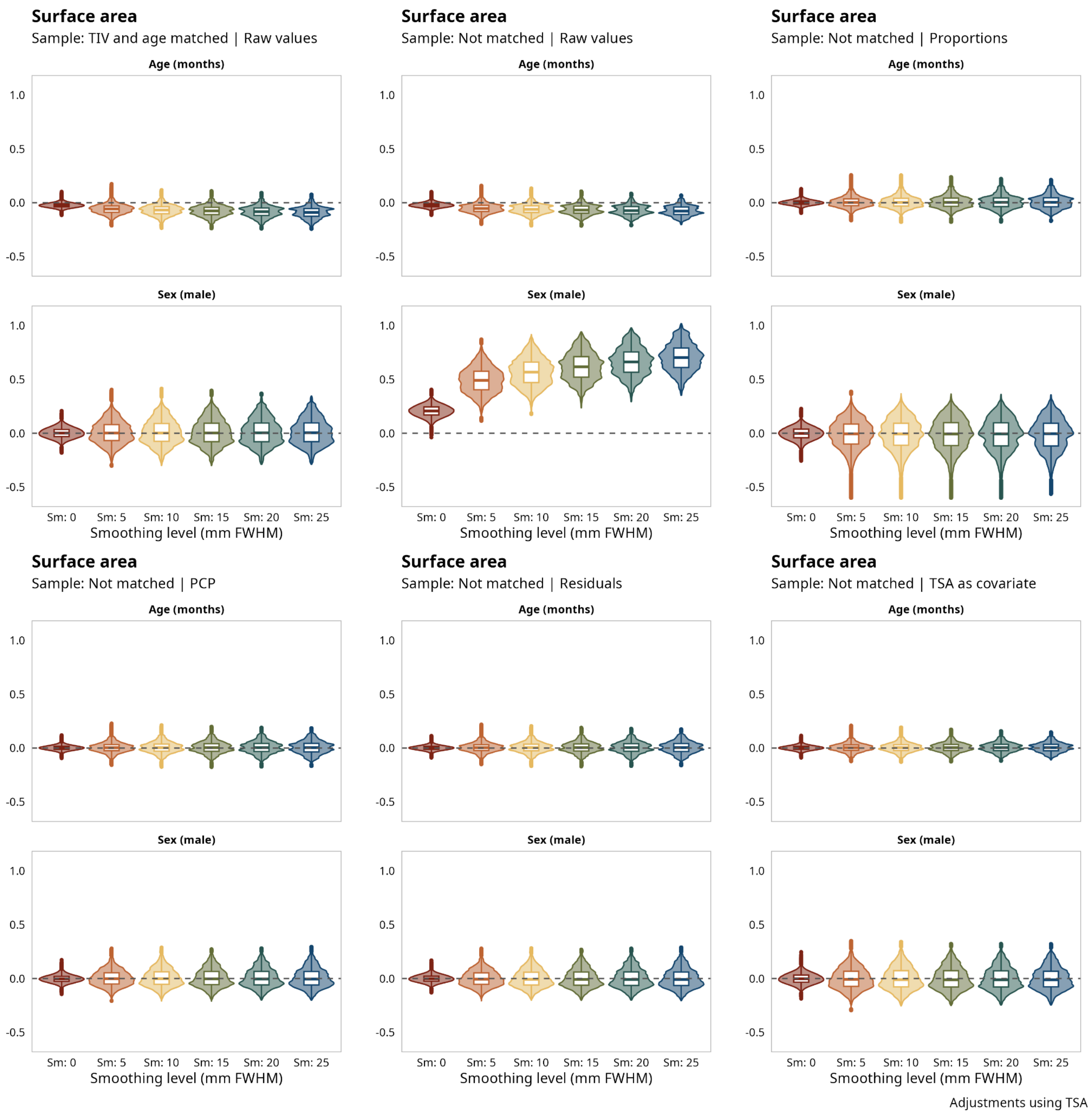


**Supplementary Figure 14.** Vertexwise Surface Area. Effect of different levels of smoothing on age and sex estimation, using the raw values in the mached and not matched samples and adjusting using total surface area (TSA) in the not matched sample.

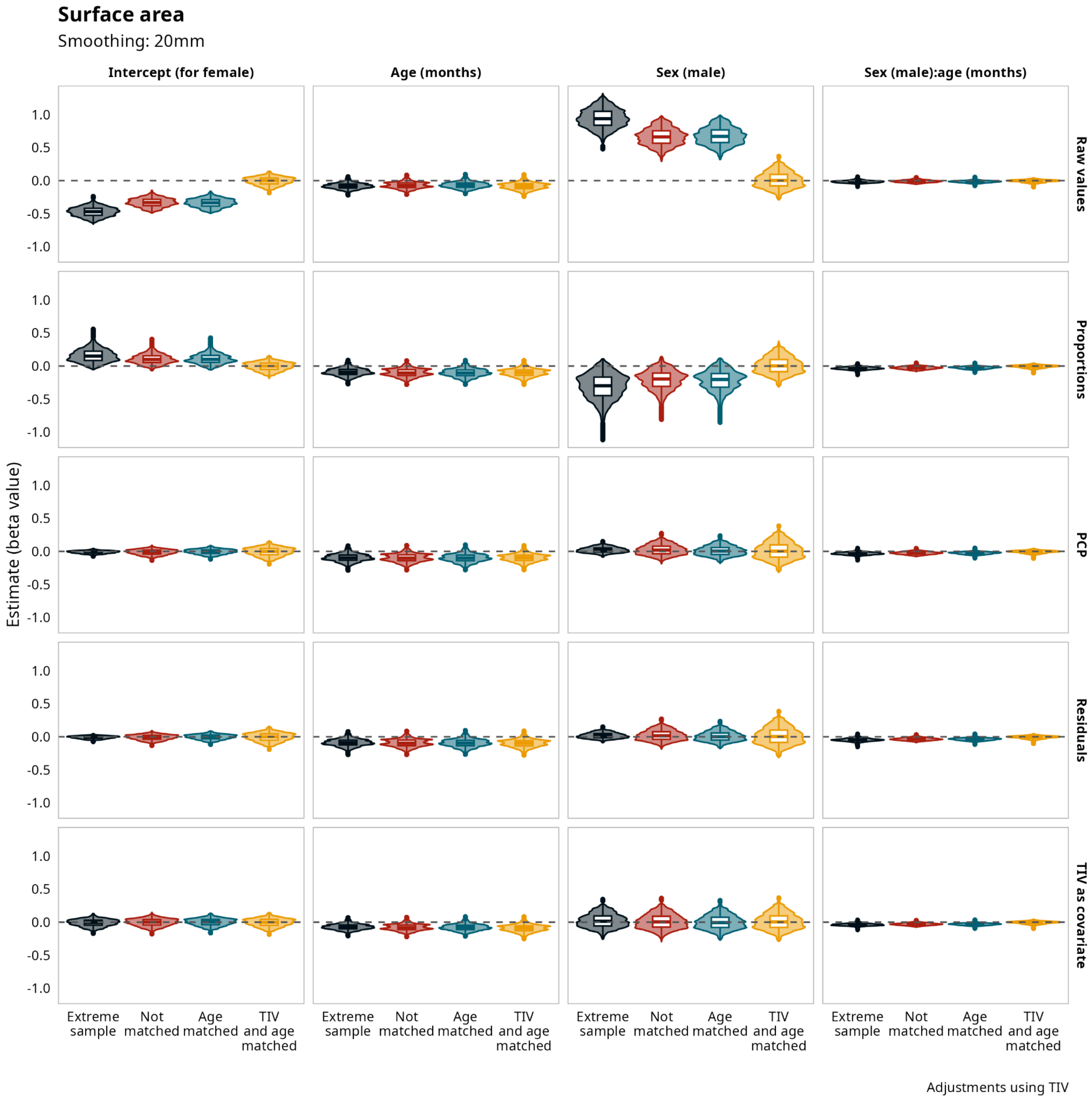


**Supplementary Figure 15.** Vertexwise Surface Area. Model's estimates for vertexwise surface area smoothed 20mm FWHM, corrected using total intracranial volume (TIV) for all the samples and all the adjustment methods.

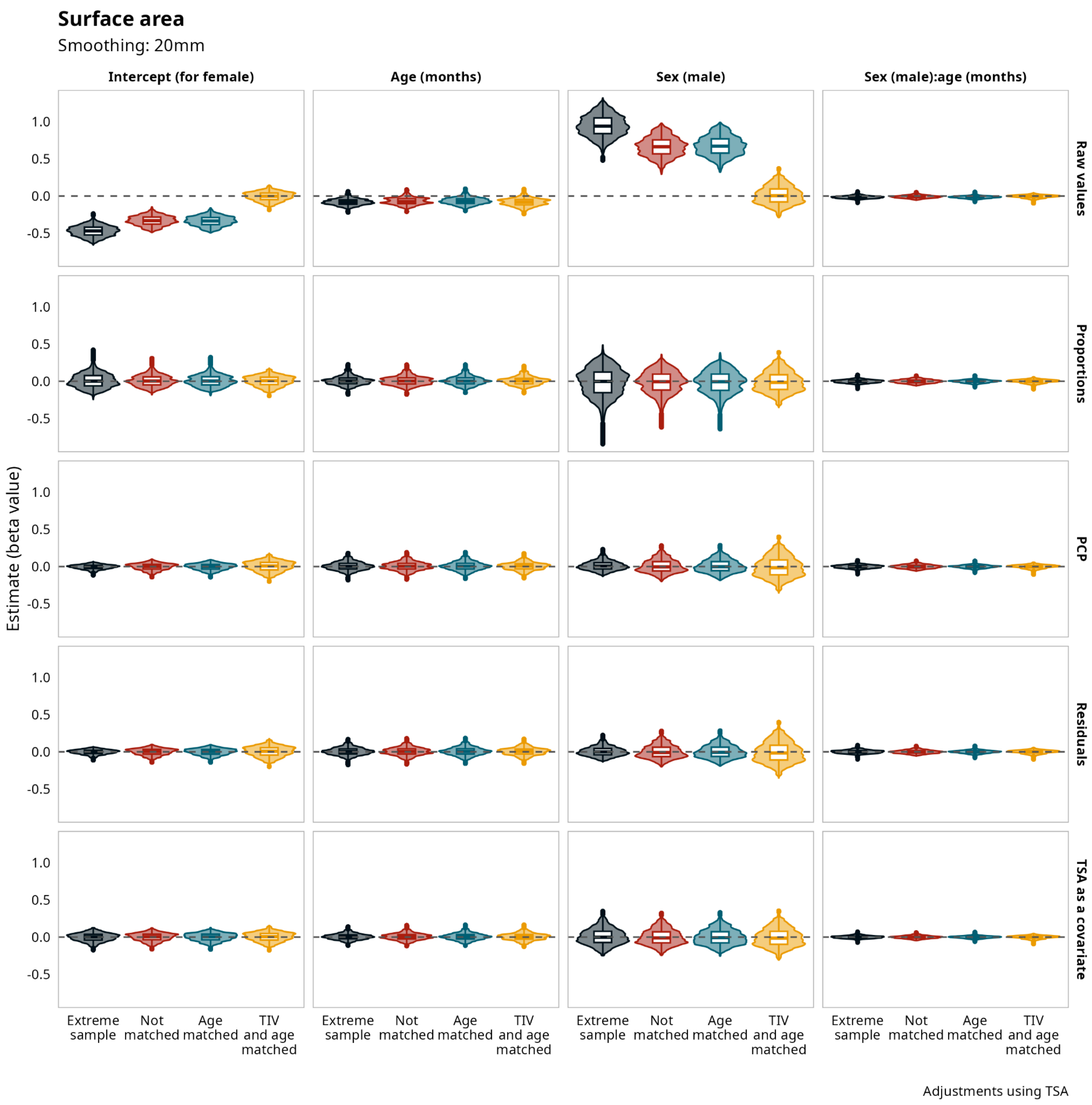


**Supplementary Figure 16.** Vertexwise Surface Area. Model's estimates for vertexwise surface area smoothed 20mm FWHM, corrected using total surface area (TSA) for all the samples and all the adjustment methods.

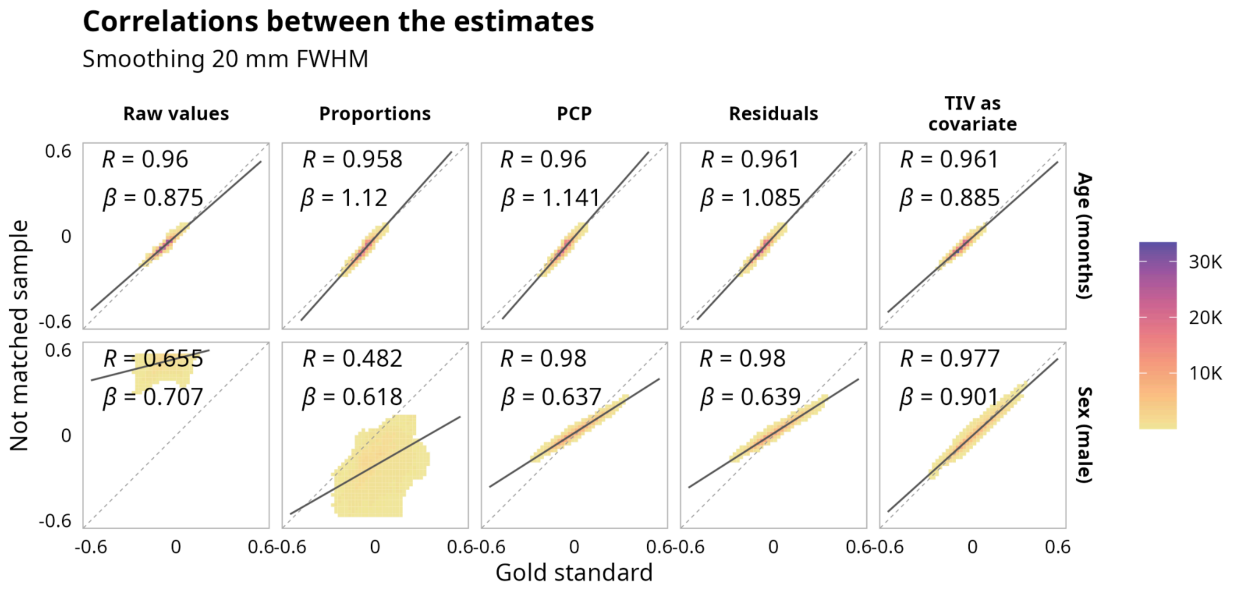


**Supplementary Figure 17.** Vertexwise Surface Area. Correlations between estimates for the matched sample without adjustment and the estimates after using different adjustment methods with total intracranial volume (TIV) and  the not matched sample, with data smoothed 20mm FWHM.

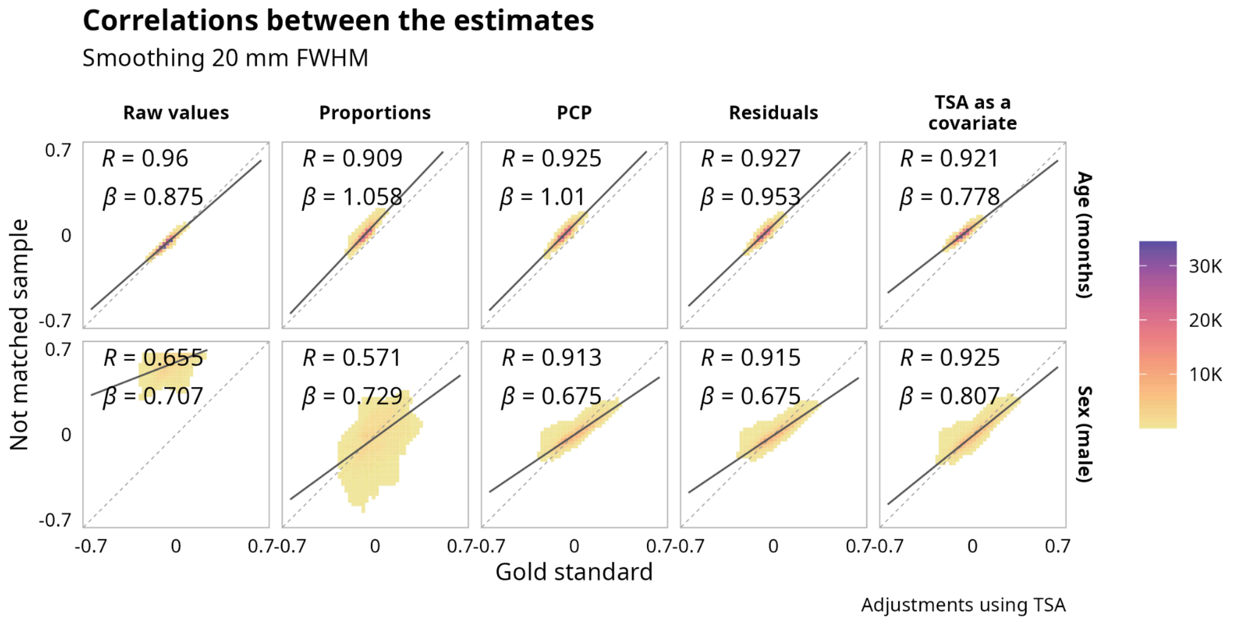


**Supplementary Figure 18.** Vertexwise Surface Area. Correlations between estimates for the matched sample without adjustment and the estimates after using different adjustment methods with total surface area (TSA) and the not matched sample, with data smoothed 20mm FWHM.

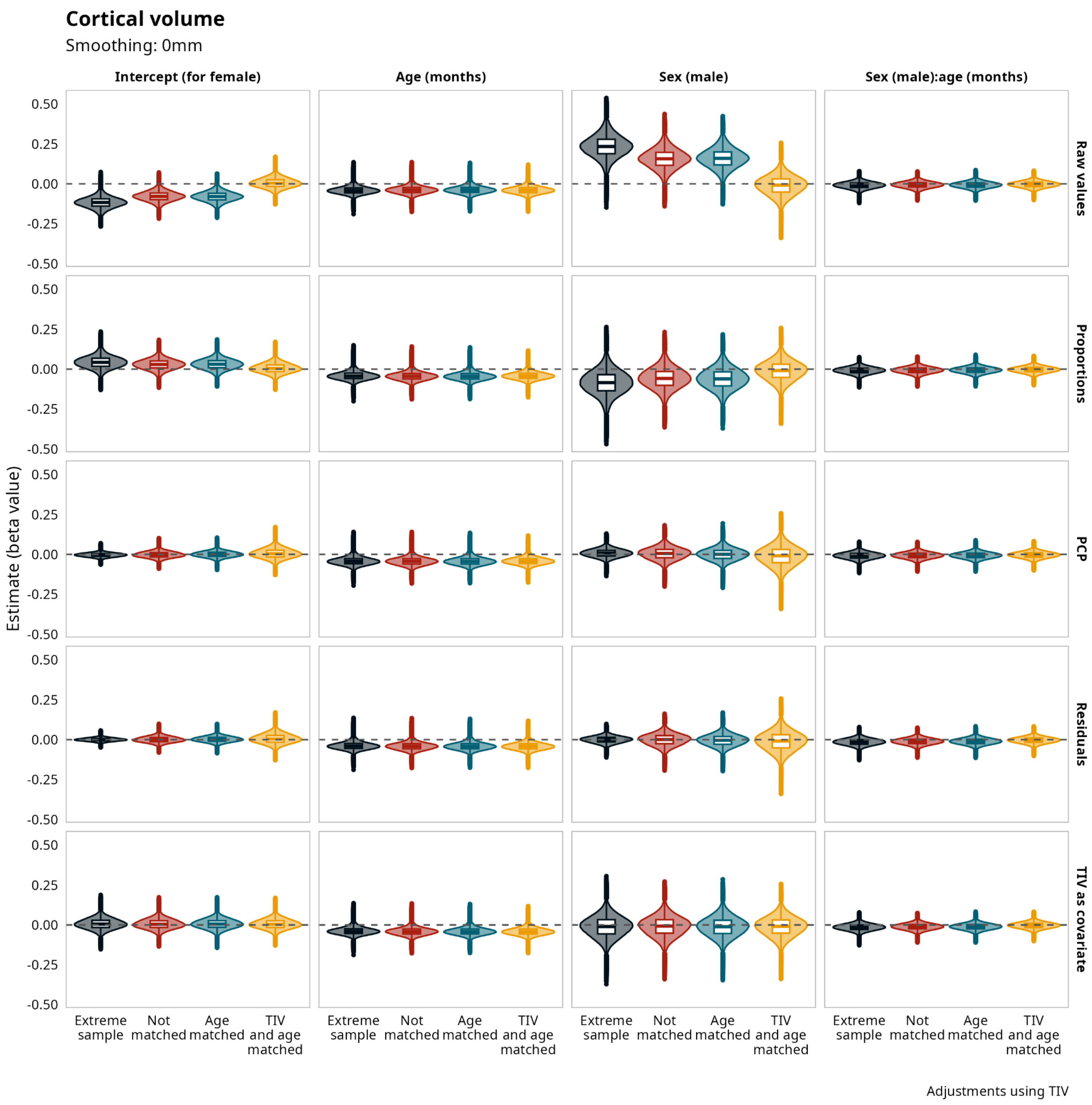


**Supplementary Figure 19.** Vertexwise Cortical Volume. Model's estimates for vertexwise cortical volume without smoothing corrected using total intracranial volume (TIV) for all the samples and all the adjustment methods.

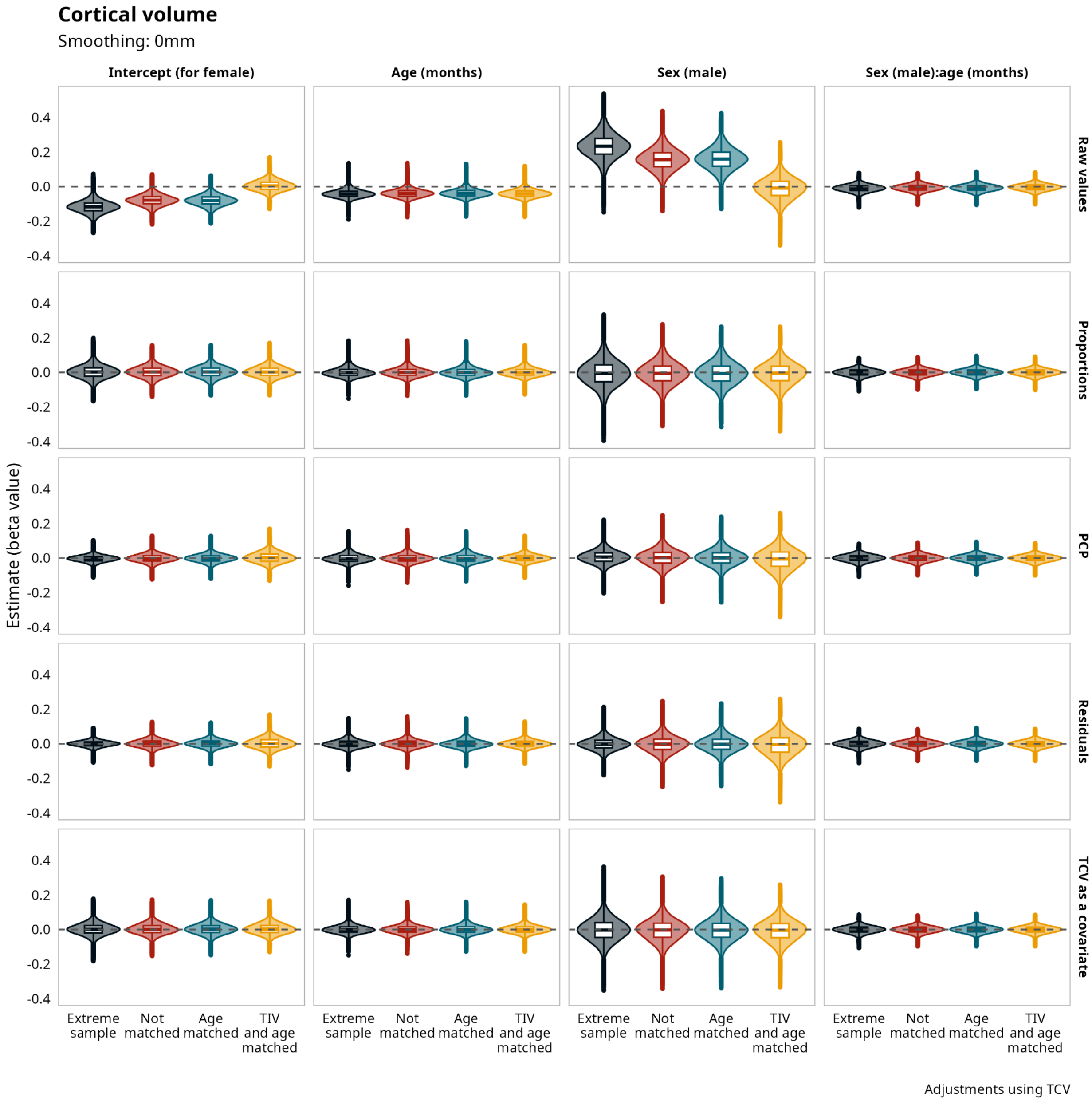


**Supplementary Figure 20. Supplementary Figure 19.** Vertexwise Cortical Volume. Model's estimates for vertexwise cortical volume without smoothing corrected using total cortical volume (TCV) for all the samples and all the adjustment methods.

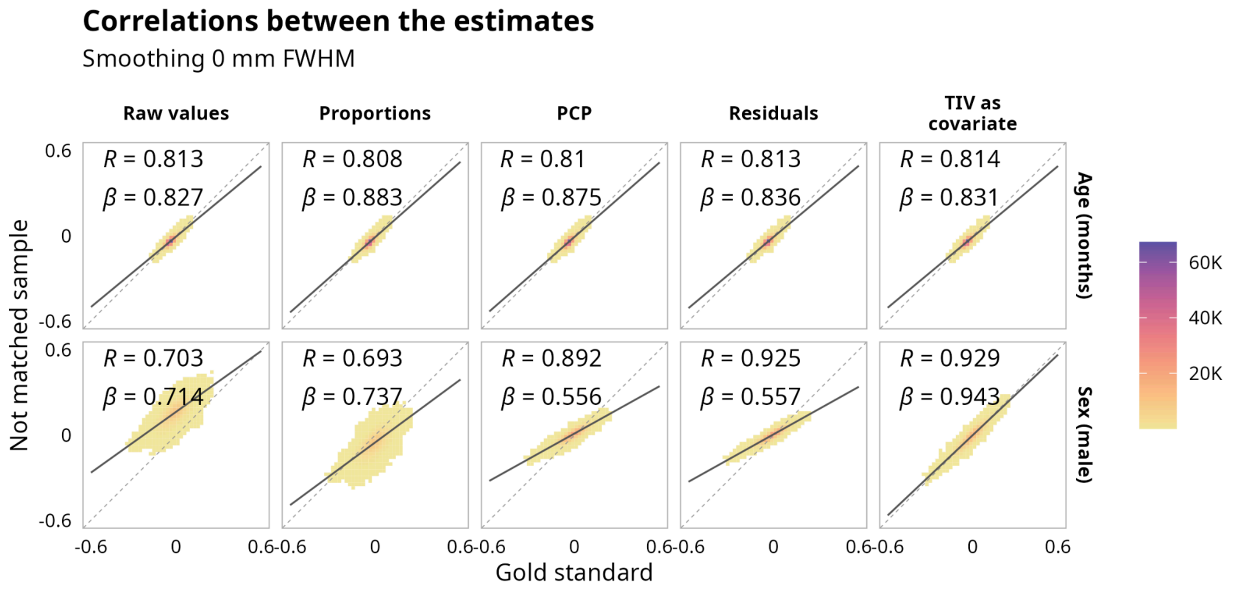


**Supplementary Figure 21.** Vertexwise Cortical Volume. Correlations between estimates for the matched sample without adjustment and the estimates after using different adjustment methods with total intracranial volume (TIV) and the not matched sample, with data without smoothing.

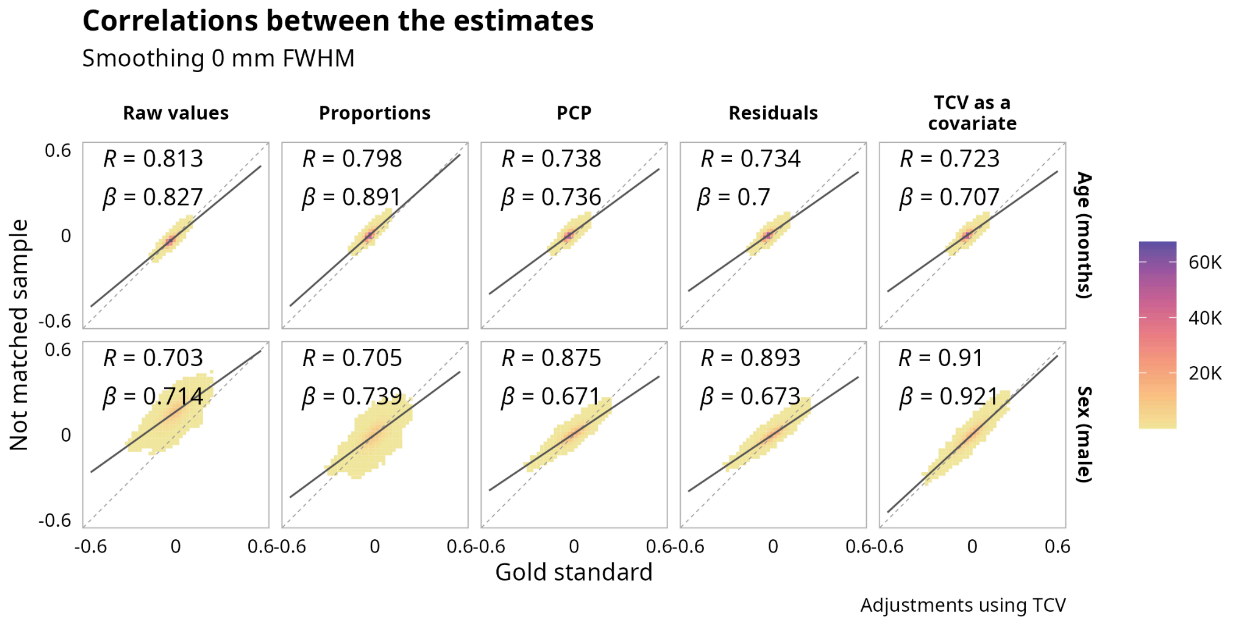


**Supplementary Figure 22.** Vertexwise Cortical Volume. Correlations between estimates for the matched sample without adjustment and the estimates after using different adjustment methods with total cortical volume (TCV) and the not matched sample, with data without smoothing.

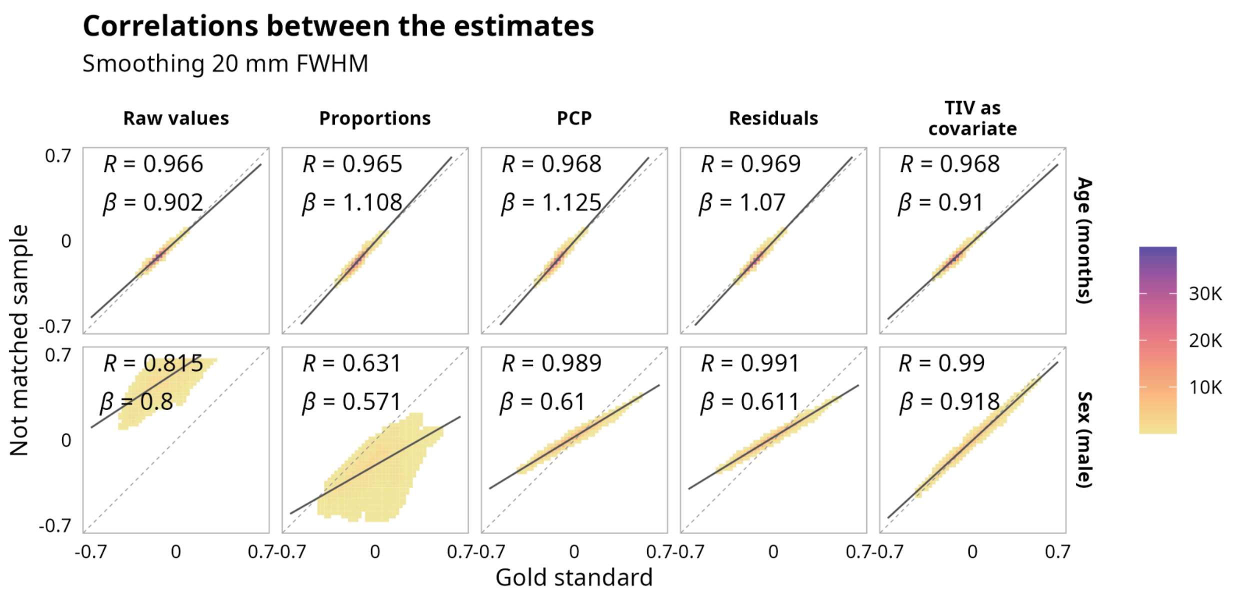


**Supplementary Figure 23.** Vertexwise Cortical Volume. Correlations between estimates for the matched sample without adjustment and the estimates after using different adjustment methods with total intracranial volume (TIV) and the not matched sample, with data smoothed at 20mm FWHM.

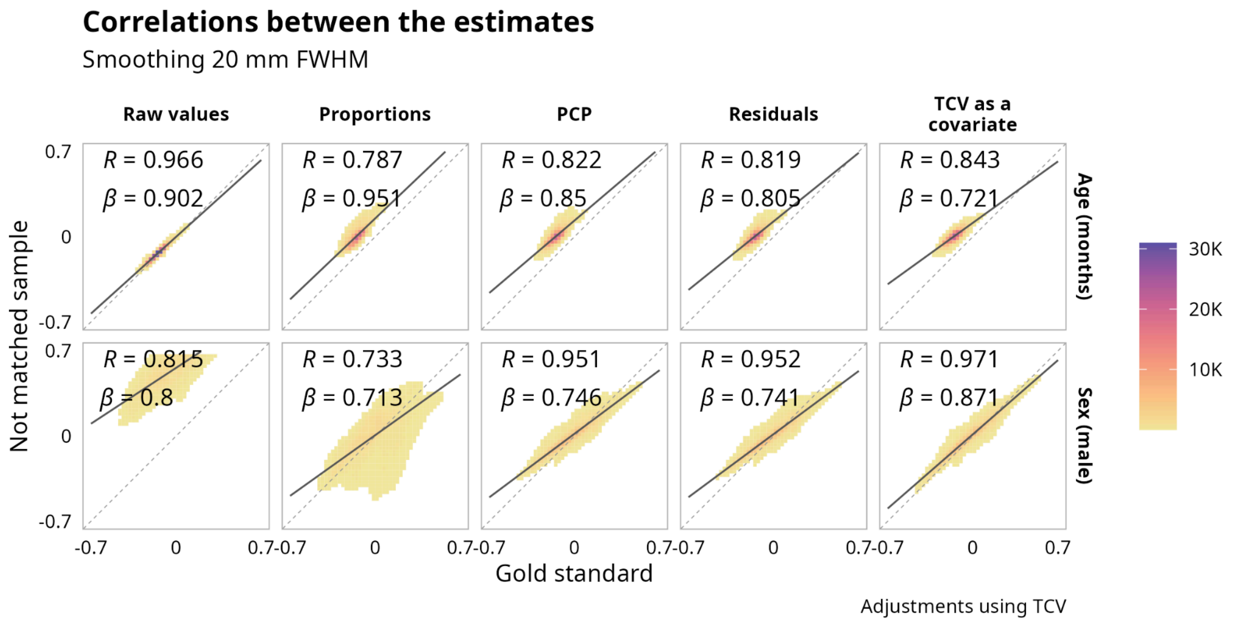


**Supplementary Figure 24.** Vertexwise Cortical Volume. Correlations between estimates for the matched sample without adjustment and the estimates after using different adjustment methods with total cortical volume (TCV) and the not matched sample, with data smoothed at 20mm FWHM.

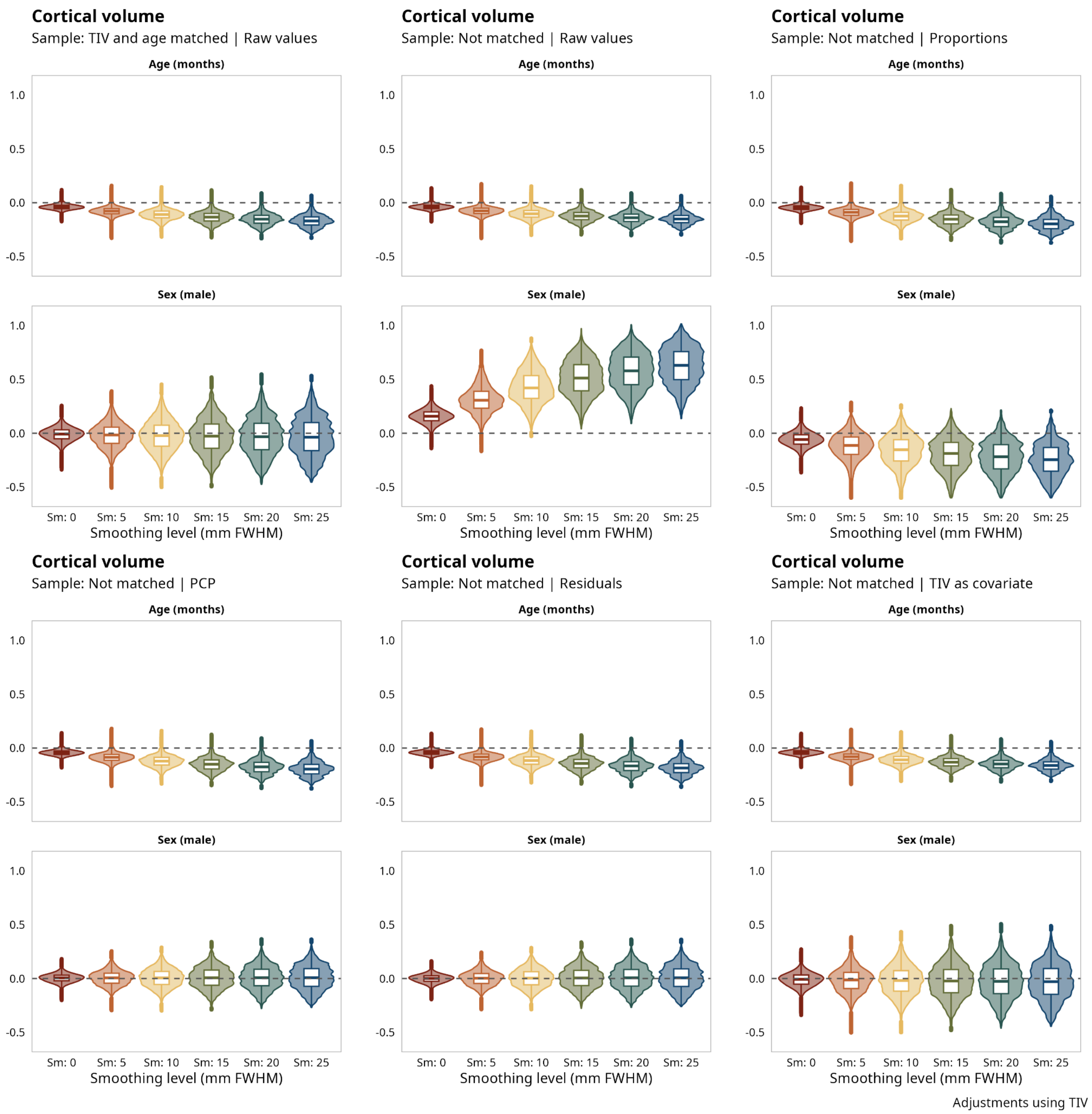


**Supplementary Figure 25.** Vertexwise Cortical Volume. Effect of different levels of smoothing on age and sex estimation, using the raw values in the mached and not matched samples and adjusting using total intracranial volume (TIV) in the not matched sample.

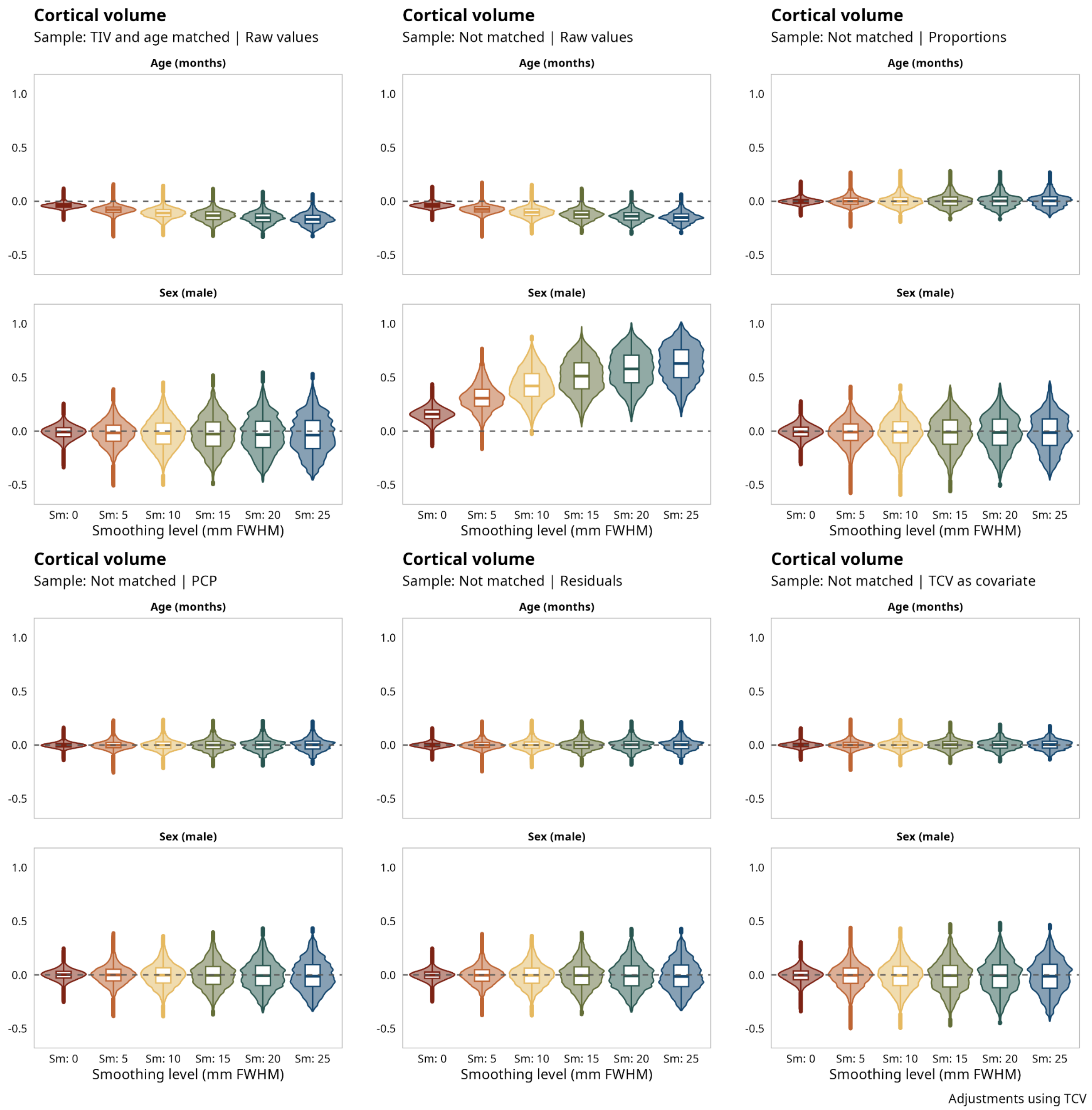


**Supplementary Figure 26.** Vertexwise Cortical Volume. Effect of different levels of smoothing on age and sex estimation, using the raw values in the mached and not matched samples and adjusting using total cortical volume (TCV) in the not matched sample.

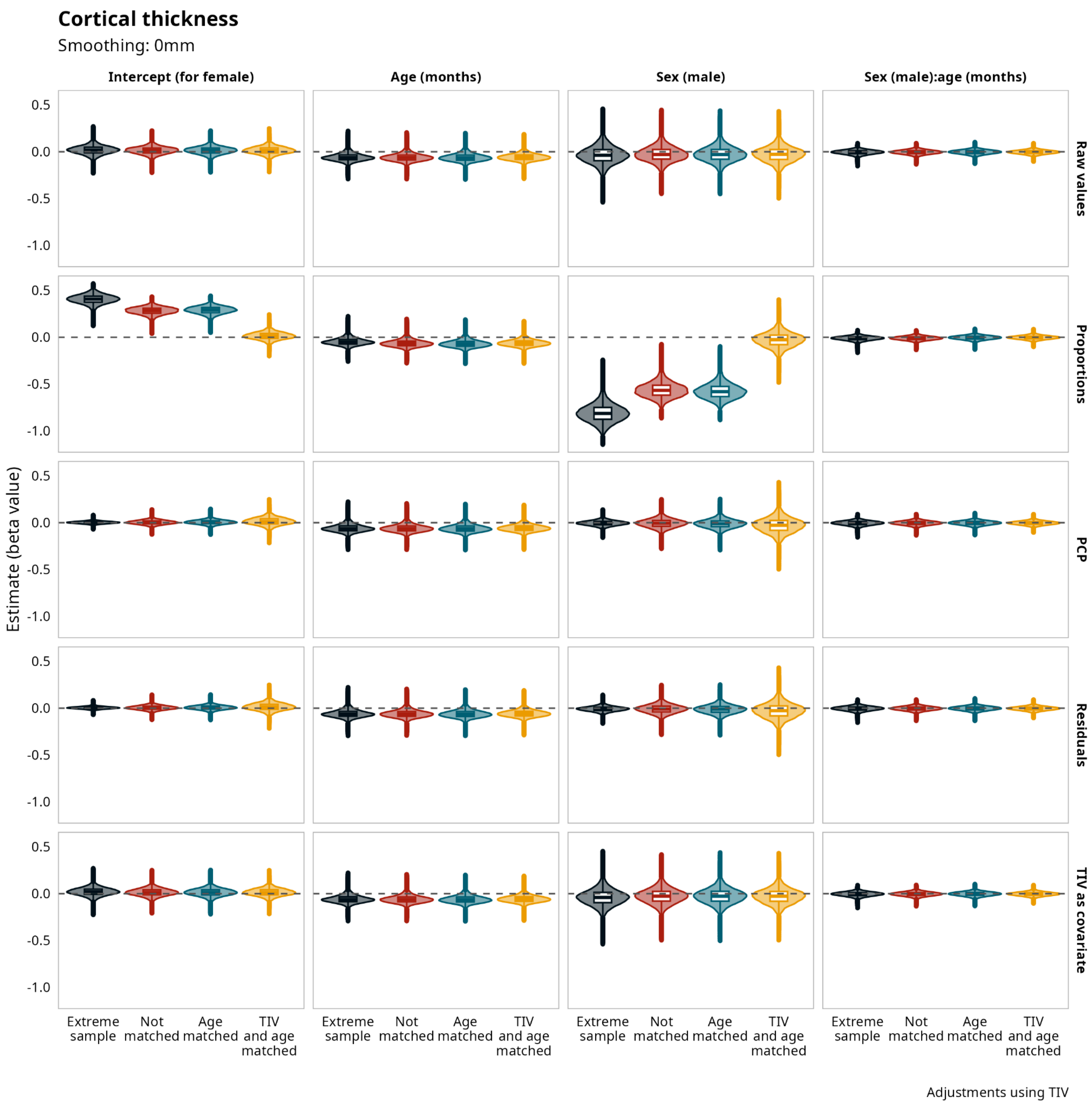


**Supplementary Figure 27.** Vertexwise Cortical Thickness. Model's estimates for vertexwise cortical thickness without smoothing corrected using total intracranial volume (TIV) for all the samples and all the adjustment methods.

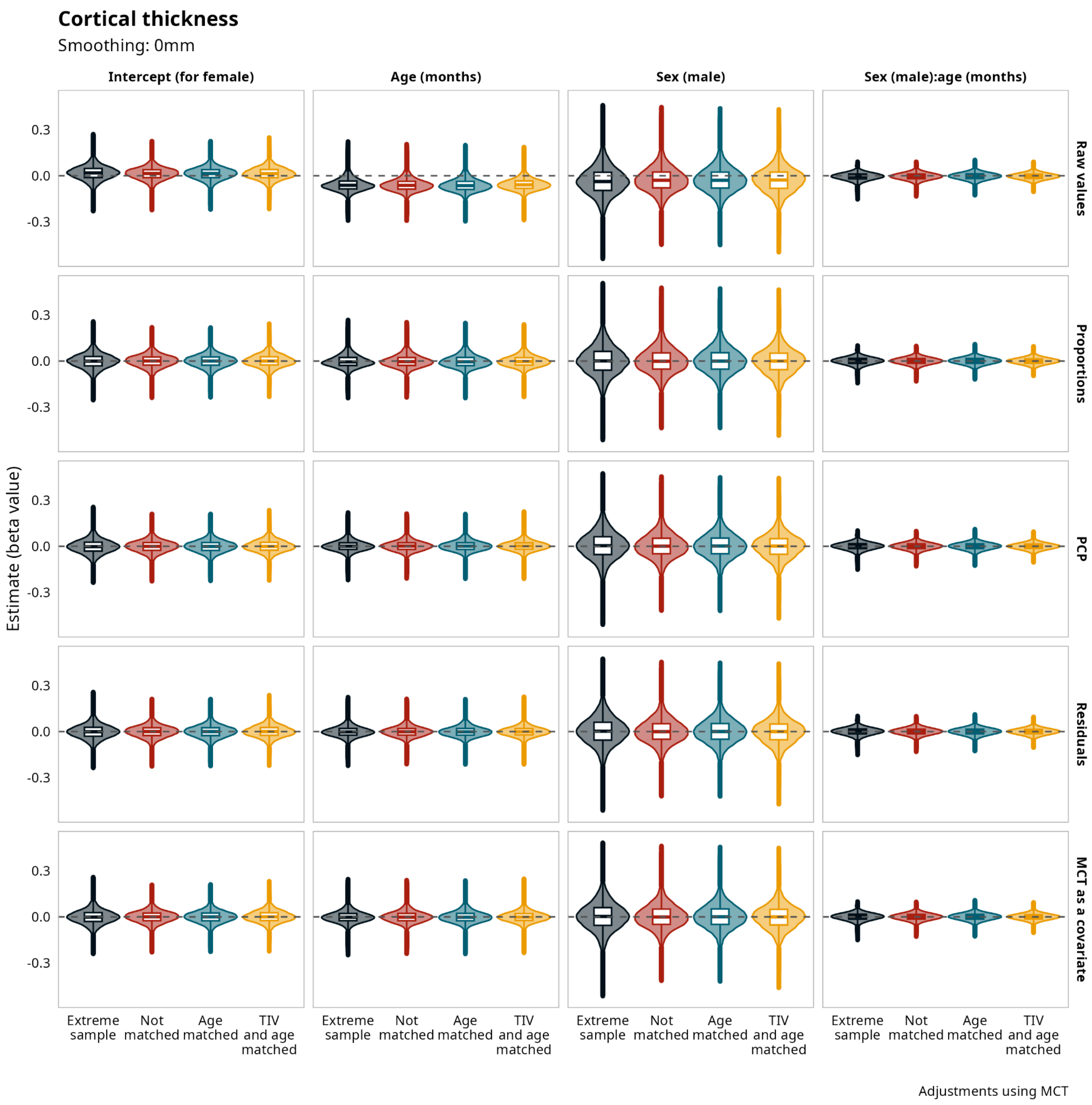


**Supplementary Figure 28.** Vertexwise Cortical Thickness. Model's estimates for vertexwise cortical thickness without smoothing corrected using mean corthical thickness (MCT) for all the samples and all the adjustment methods.

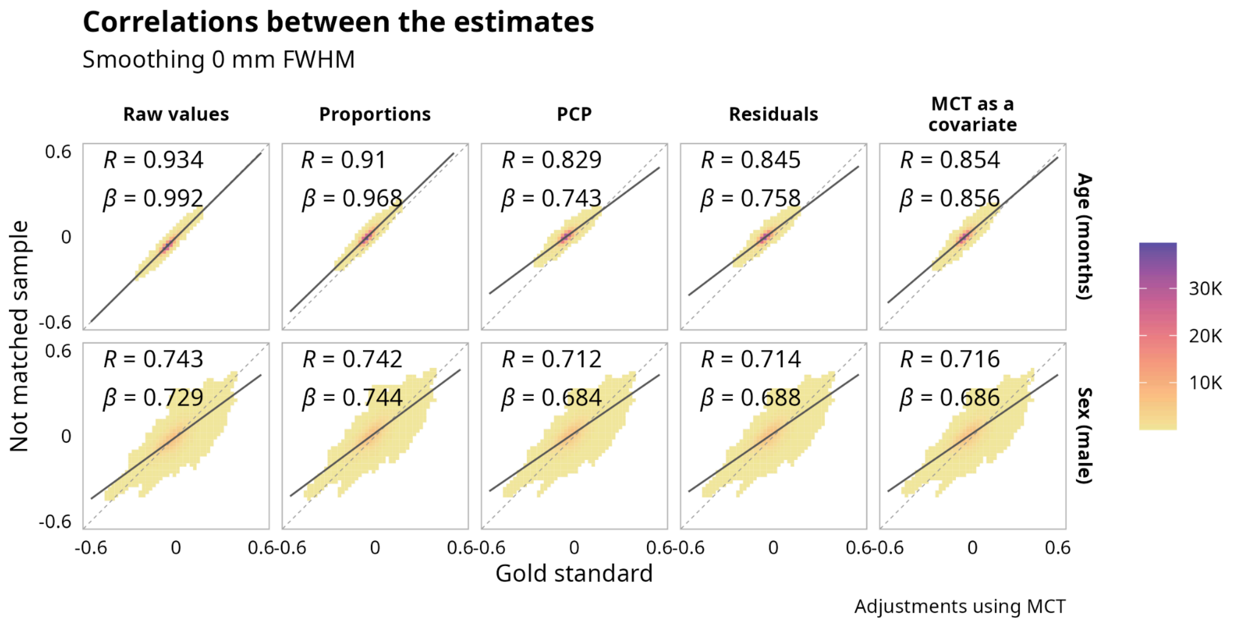


**Supplementary Figure 29.** Vertexwise Cortical Thickness. Correlations between estimates for the matched sample without adjustment and the estimates after using different adjustment methods with mean cortical thickness (MCT) and the not matched sample, with data without smoothing.

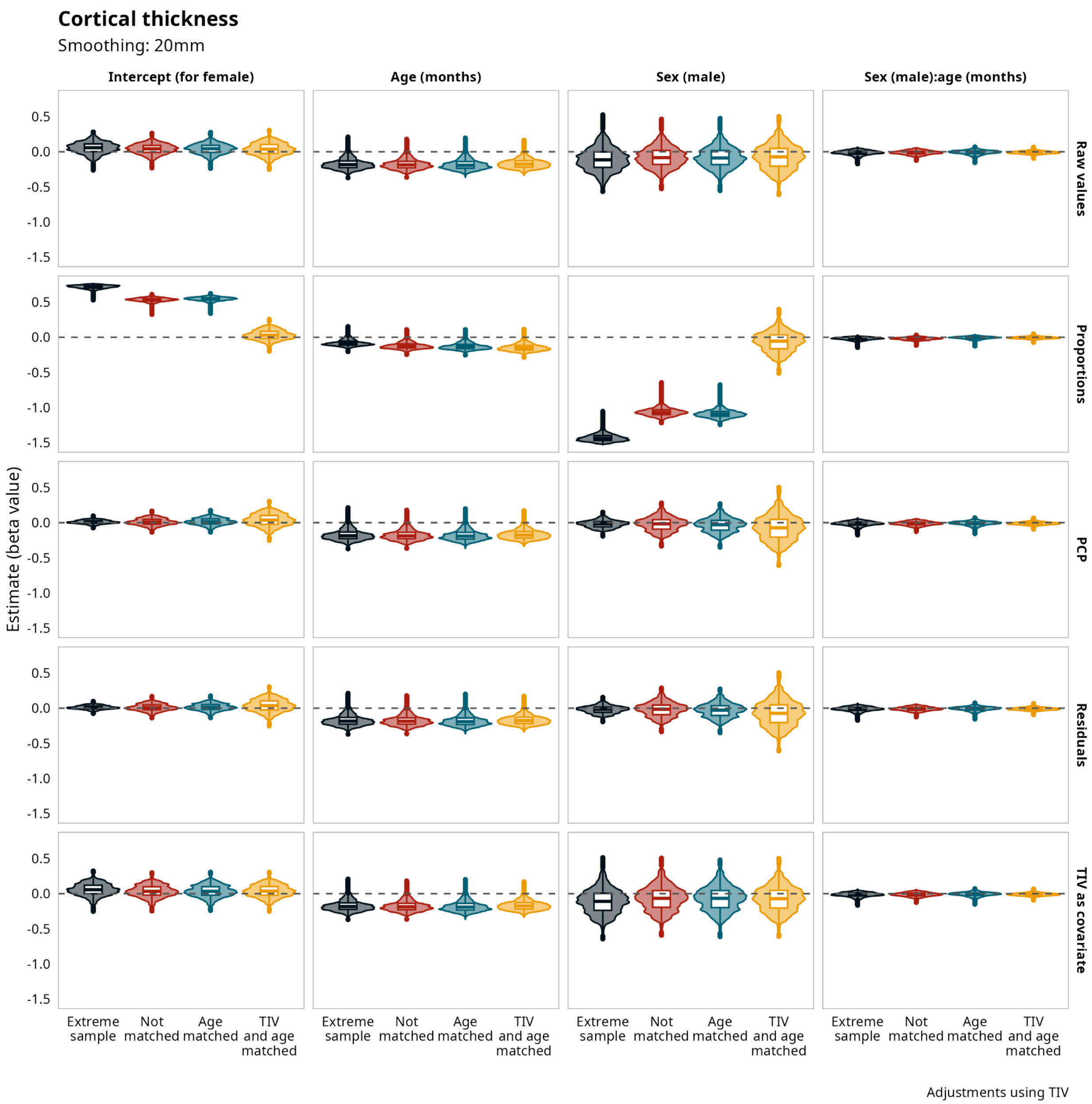


**Supplementary Figure 30.** Vertexwise Cortical Thickness. Model's estimates for vertexwise cortical thickness smoothed at 20mm FWHM corrected using total intracranial volume (TIV)  for all the samples and all the adjustment methods.

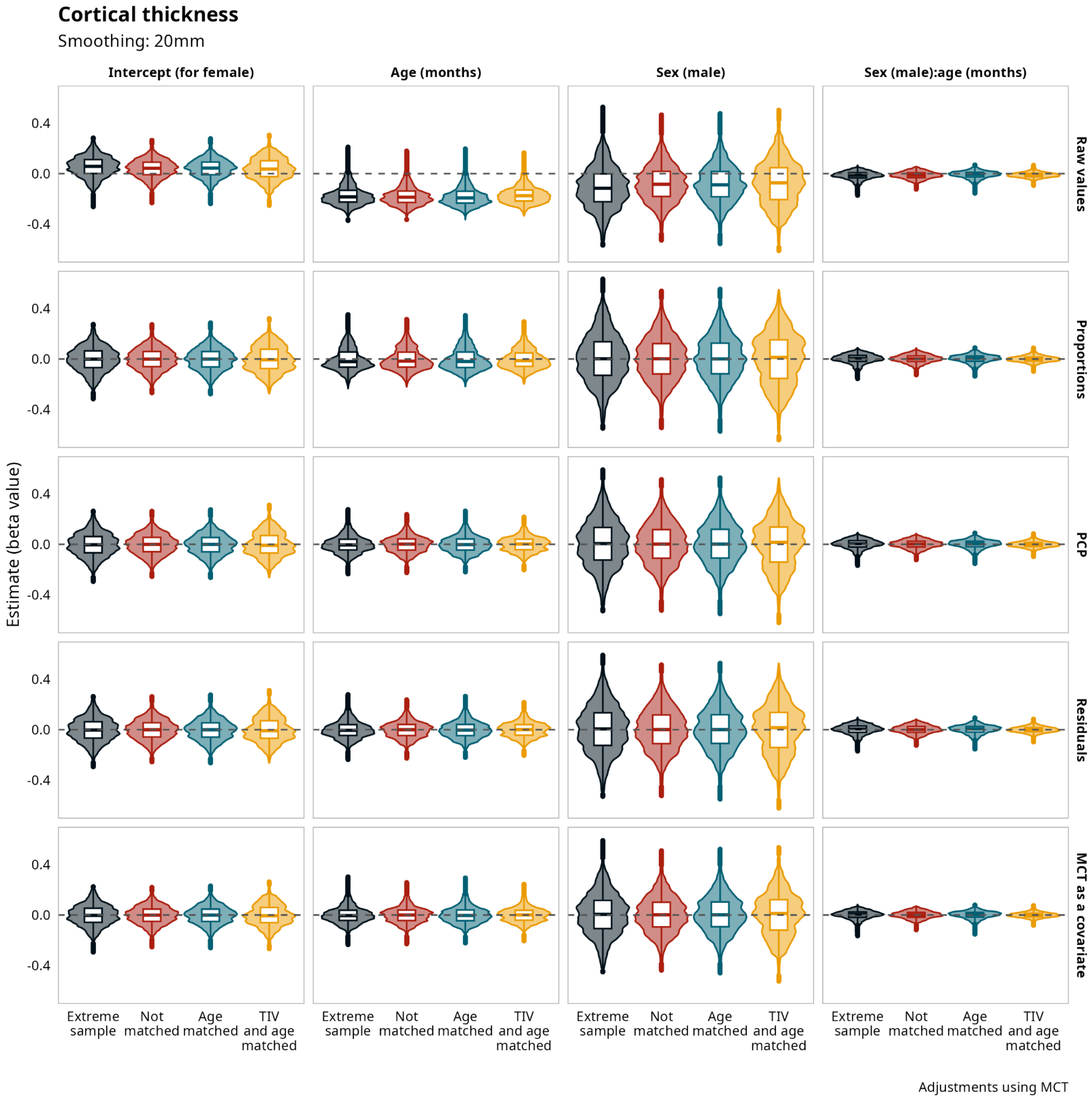


**Supplementary Figure 31.** Vertexwise Cortical Thickness. Model's estimates for vertexwise cortical thickness smoothed at 20mm FWHM corrected using mean cortical thickness (MCT) for all the samples and all the adjustment methods.

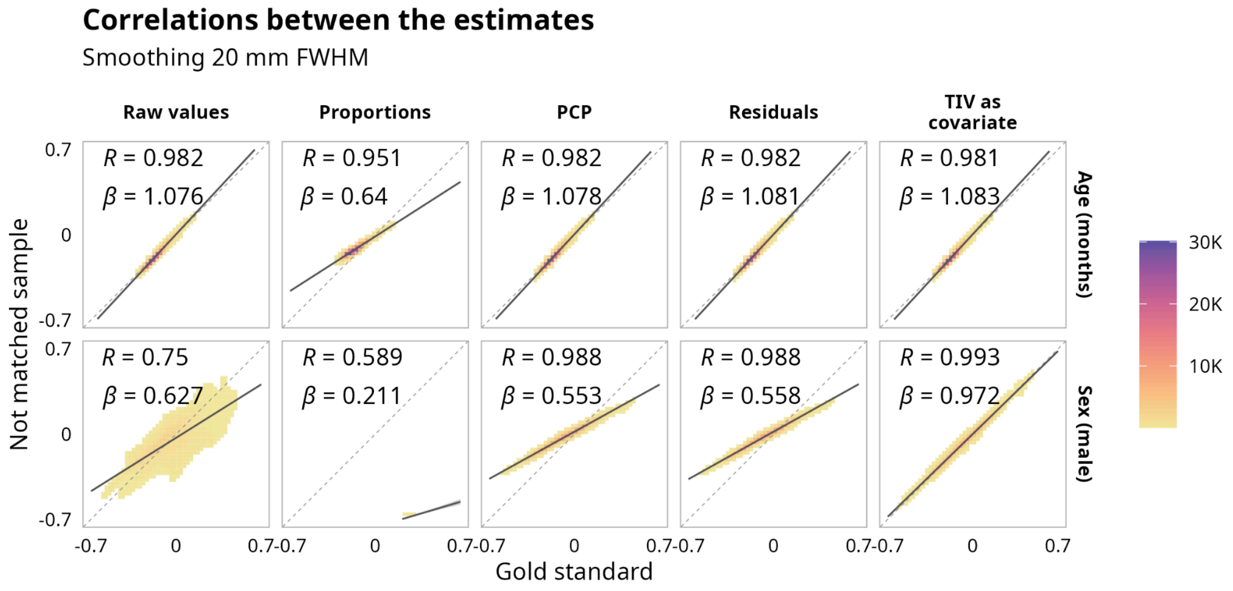


**Supplementary Figure 33.** Vertexwise Cortical Thickness. Correlations between estimates for the matched sample without adjustment and the estimates after using different adjustment methods with total intracranial volume (TIV) and the not matched sample, with data smoothed 20mm FWHM.

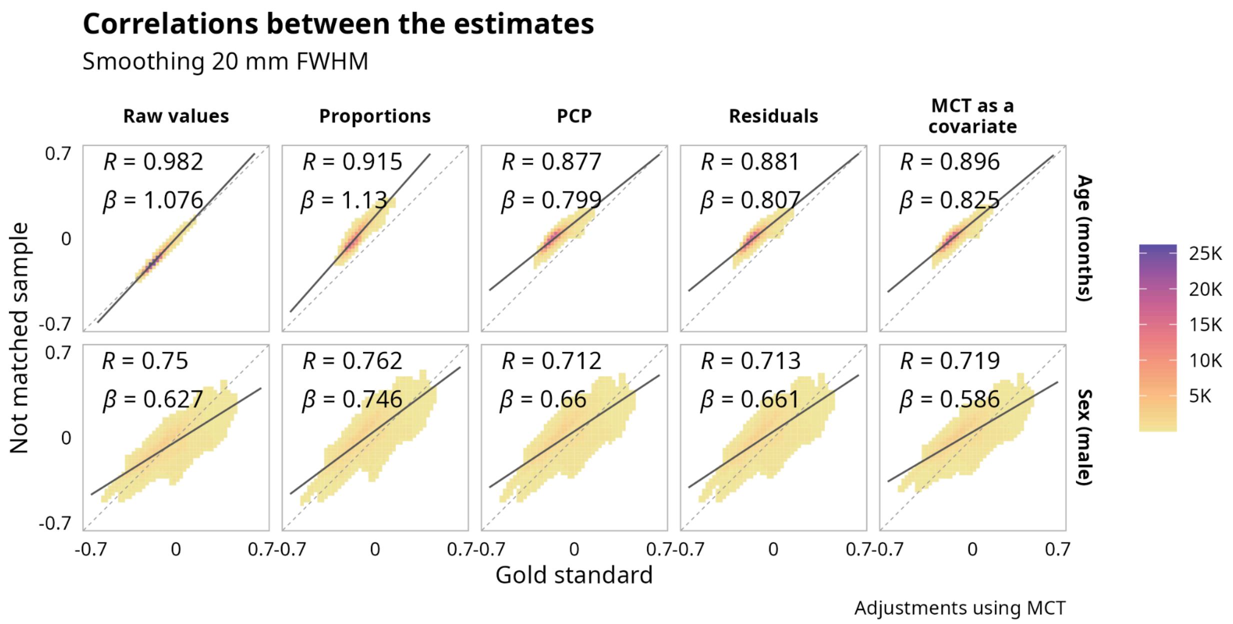


**Supplementary Figure 34.** Vertexwise Cortical Thickness. Correlations between estimates for the matched sample without adjustment and the estimates after using different adjustment methods with mean cortical thickness (MCT) and the not matched sample, with data smoothed 20mm FWHM.

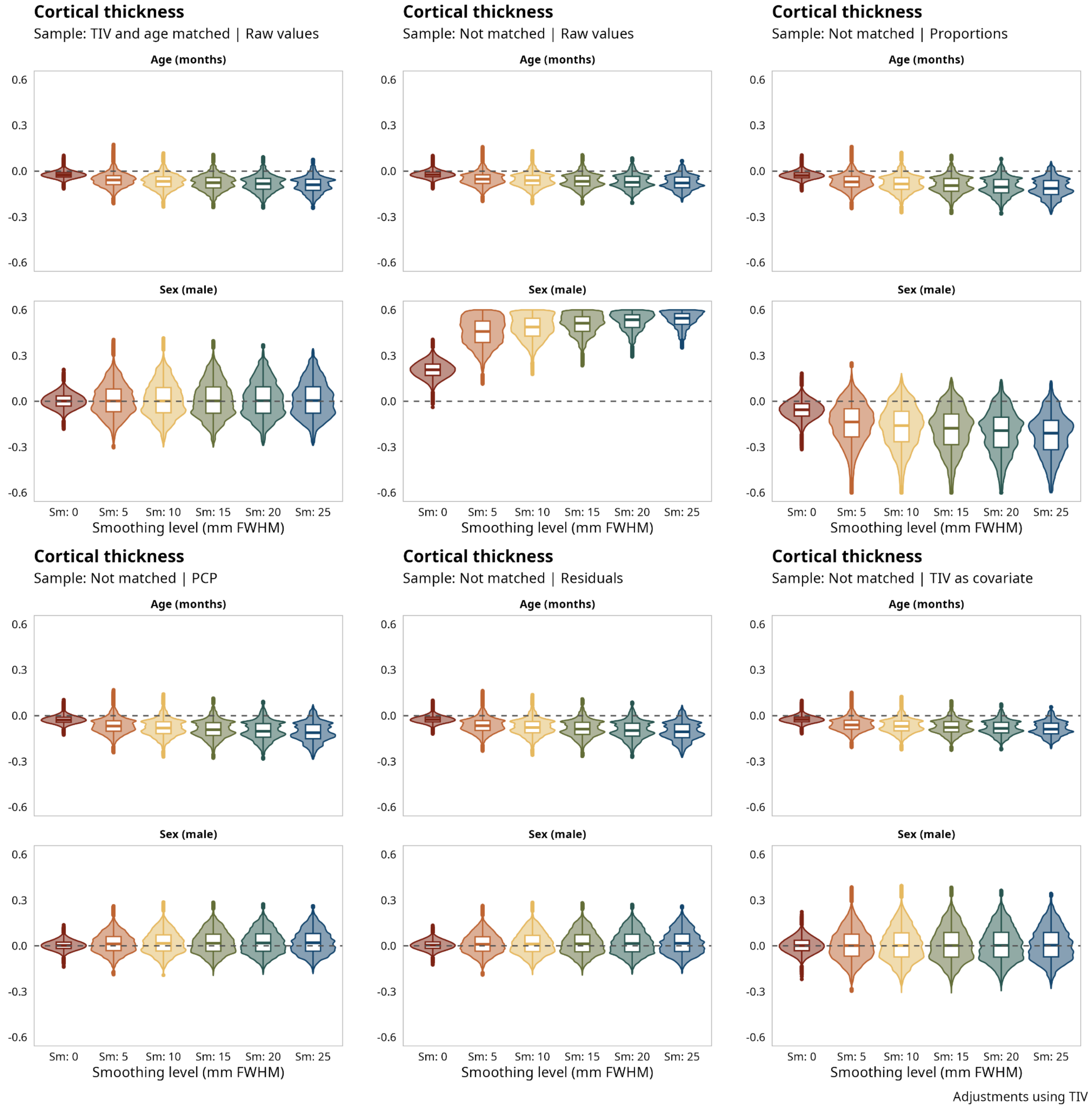


**Supplementary Figure 35.** Vertexwise Cortical Thickness. Effect of different levels of smoothing on age and sex estimation, using the raw values in the mached and not matched samples and adjusting using total intracranial volume (TIV) in the not matched sample.

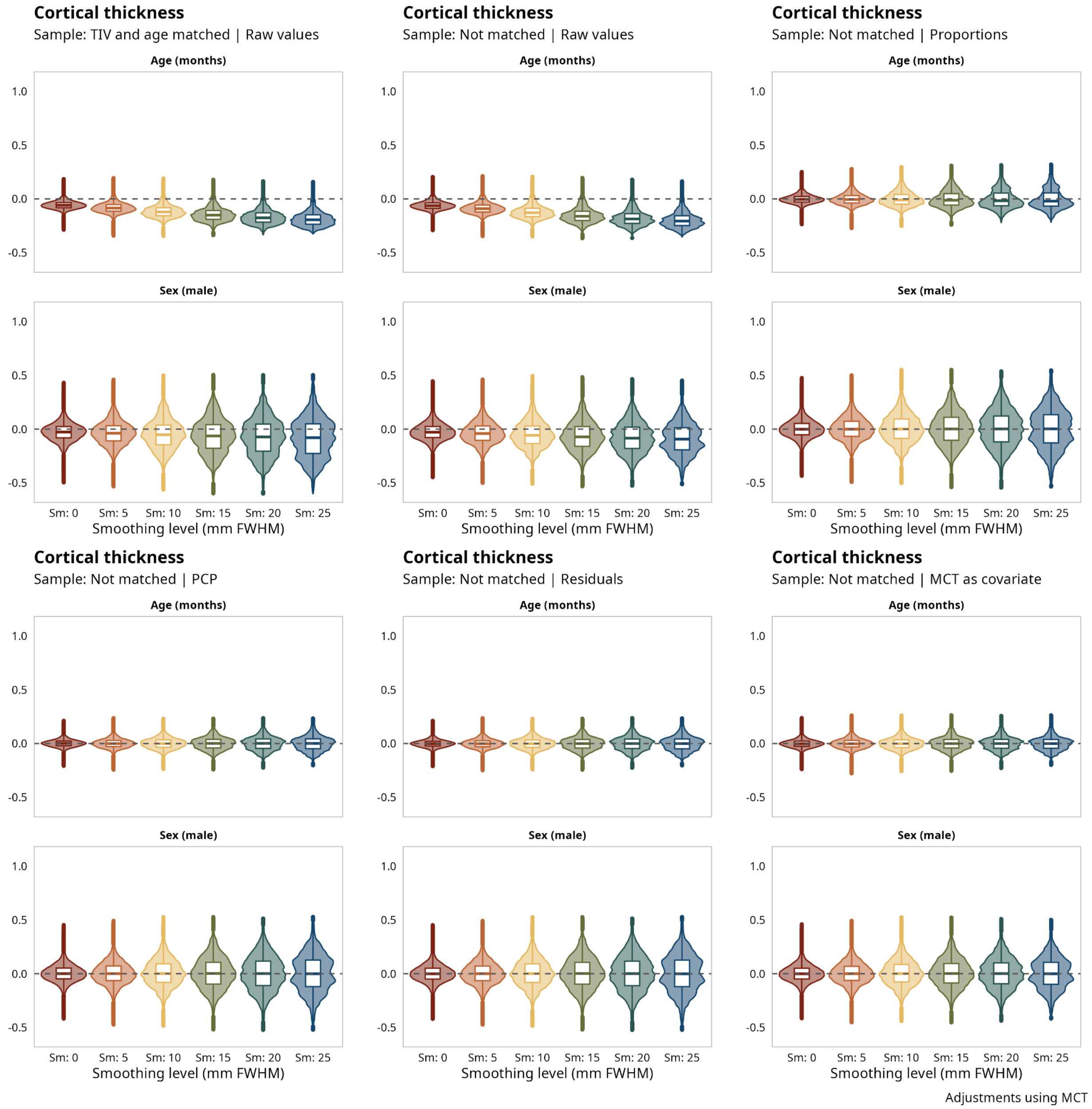


**Supplementary Figure 36.** Vertexwise Cortical Thickness. Effect of different levels of smoothing on age and sex estimation, using the raw values in the mached and not matched samples and adjusting using mean cortical thickness (MCT) in the not matched sample.

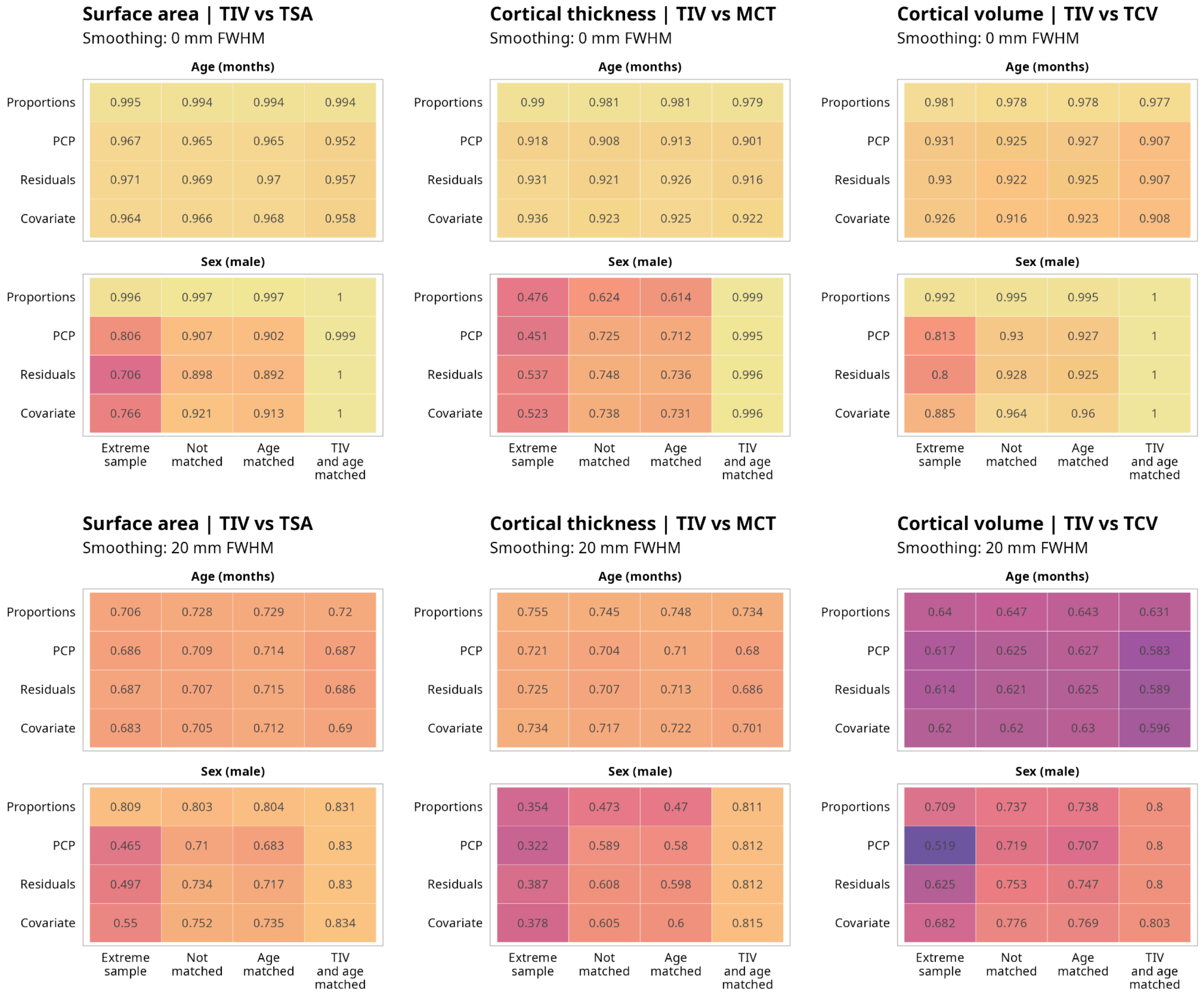


**Supplementary Figure 37.** Correlations between the estimates using different global measures for adjustment.

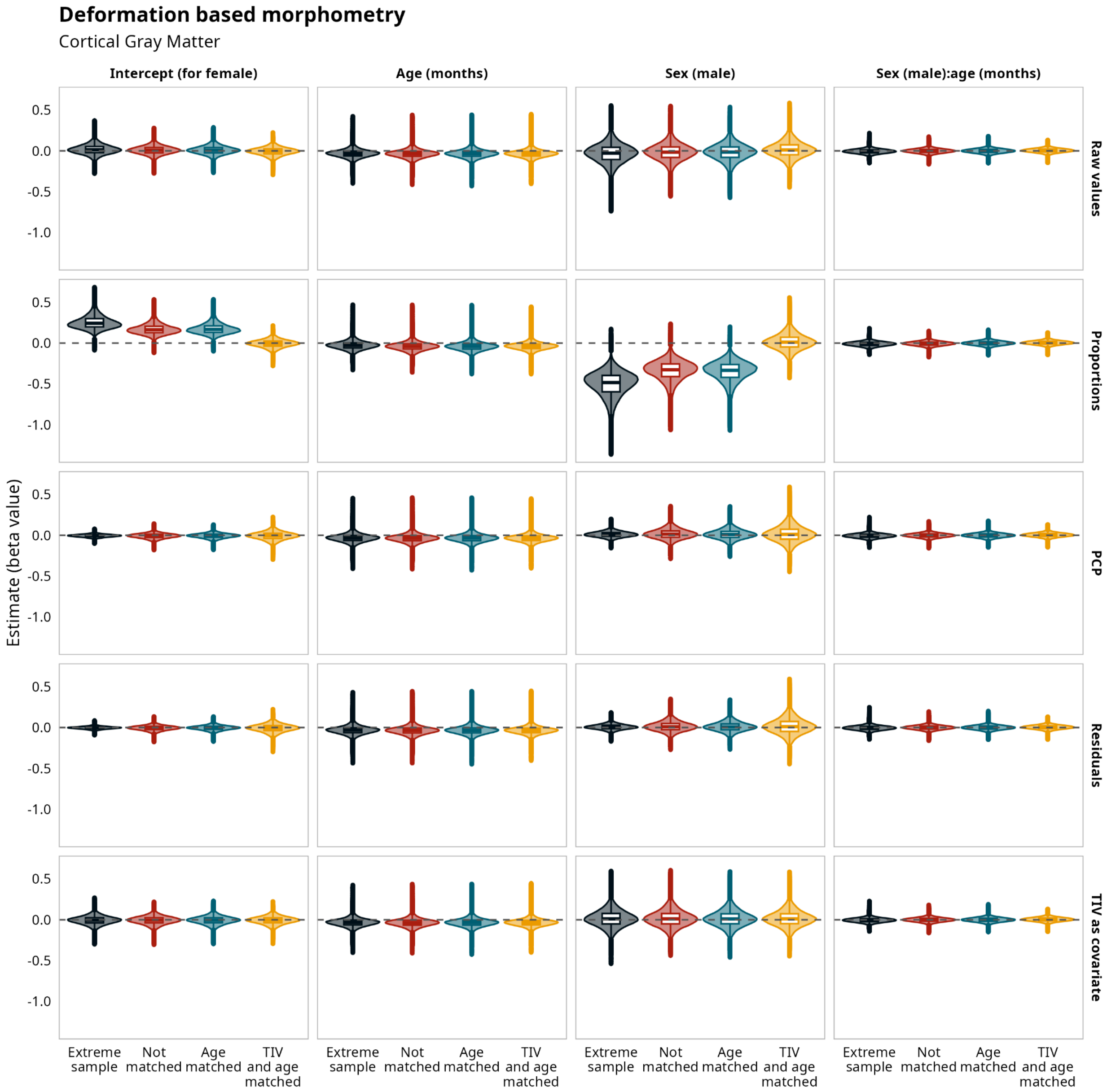


**Supplementary Figure 38.** Voxelwise deformation based morphometry. Model's estimates for voxelwise deformation based morphometry for the cortical gray matter for all the samples and all the adjustment methods.

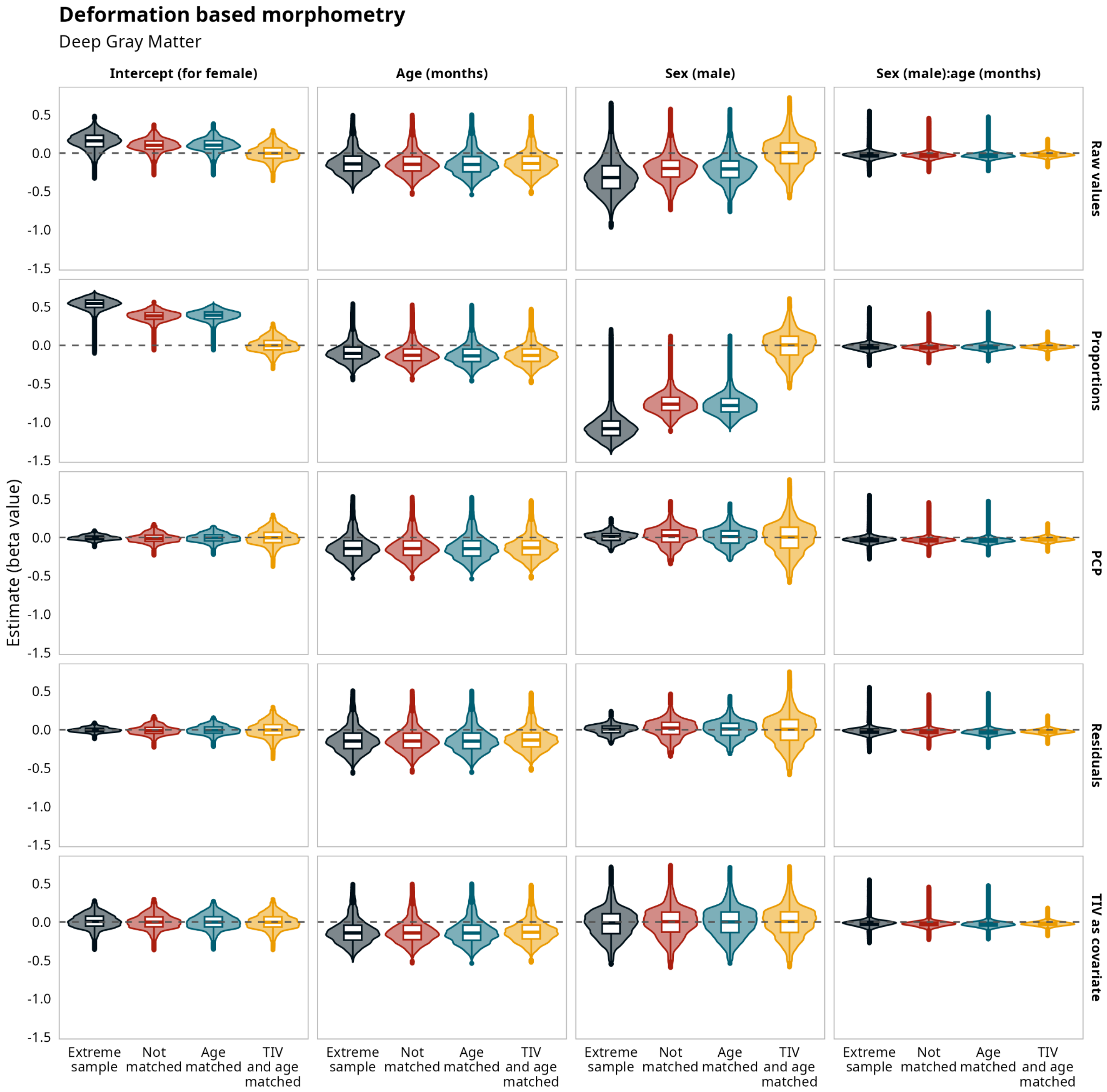


**Supplementary Figure 39.** Voxelwise deformation based morphometry. Model's estimates for voxelwise deformation based morphometry for the deep gray matter for all the samples and all the adjustment methods.

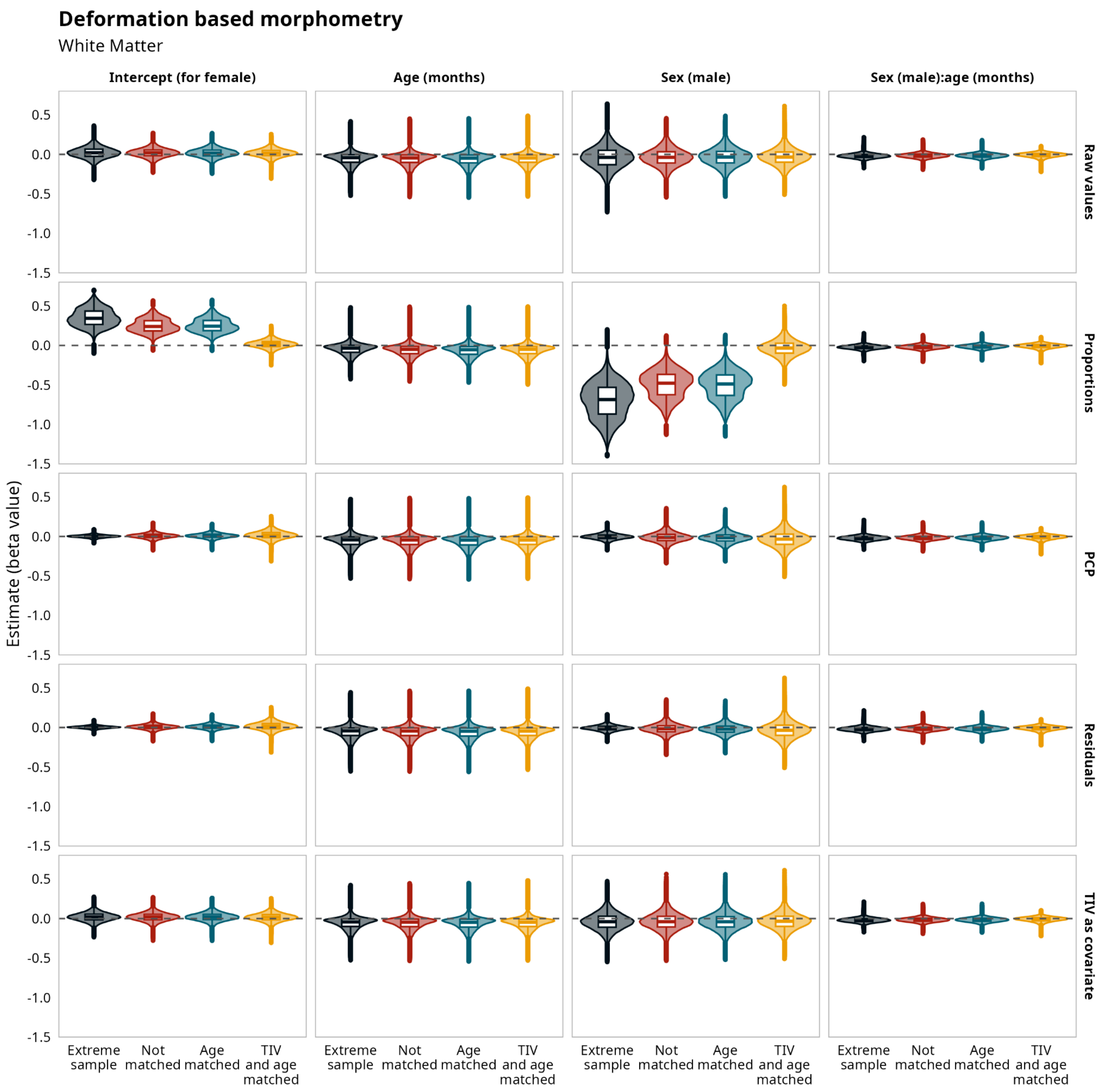


**Supplementary Figure 40.** Voxelwise deformation based morphometry. Model's estimates for voxelwise deformation based morphometry for the white matter for all the samples and all the adjustment methods.

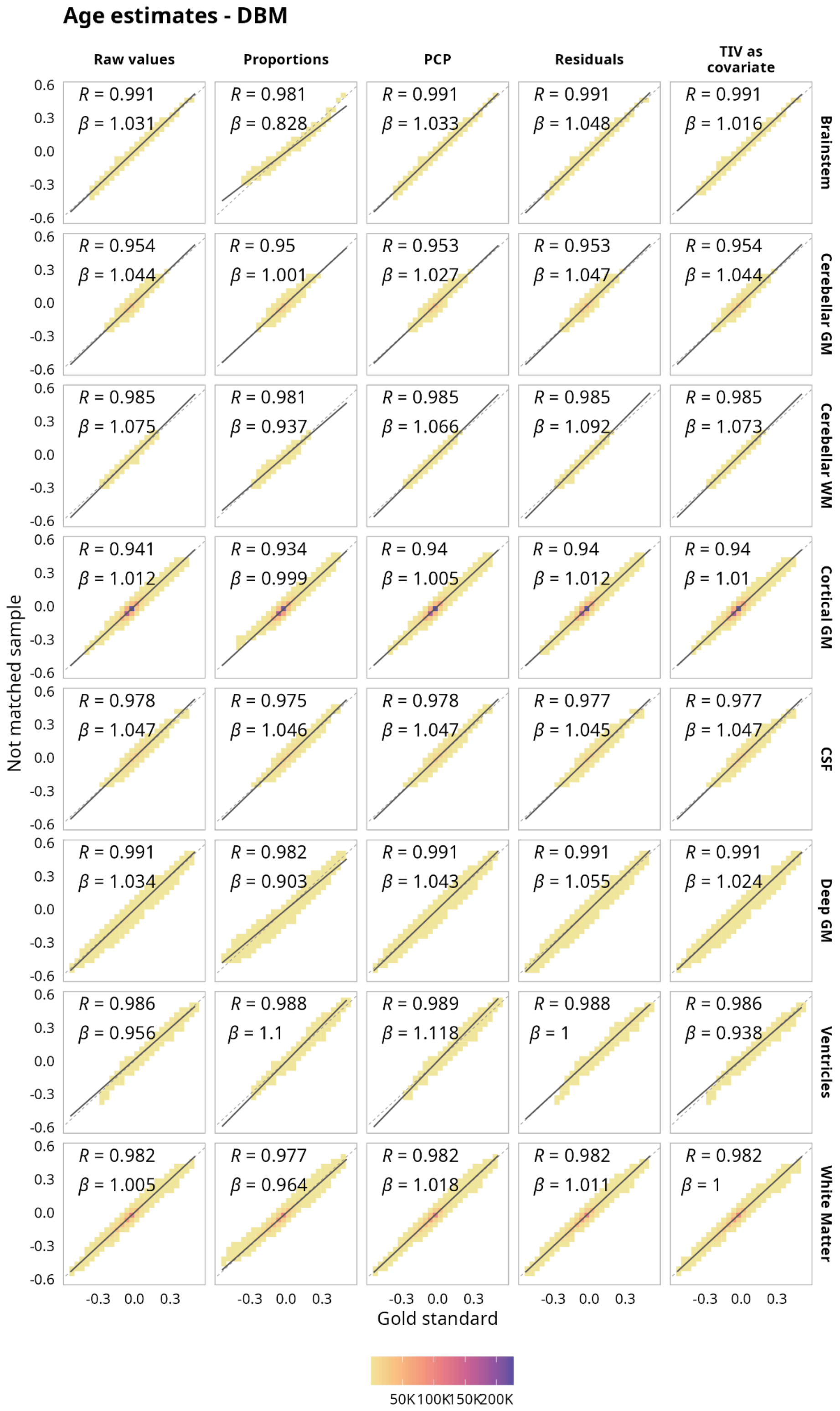


**Supplementary Figure 41.** Voxelwise Deformation Based Morphometry. Correlations between the age estimates for the matched sample without adjustment and the estimates after using different adjustment methods for the not matched sample on Voxelwise Deformation Based Morphometry across different tissue types.
